# Accurate Screening for Early-Stage Breast Cancer by Detection and Profiling of Circulating Tumor Cells

**DOI:** 10.1101/2022.05.10.22274886

**Authors:** Timothy Crook, Darshana Patil, Dadasaheb Akolkar, Anantbhushan Ranade, Sewanti Limaye, Raymond Page, Vineet Datta, Pradip Fulmali, Sachin Apurwa, Stefan Schuster, Ajay Srinivasan, Rajan Datar

## Abstract

**BACKGROUND:** Screening of asymptomatic women for early detection of Breast Cancer (BrC) is associated with improved survival. Presently, mammography is the standard of care (SoC) for BrC screening but has lower accuracy for invasive cancers as well as in women with higher breast tissue density.

**METHODS:** In this manuscript, we describe an accurate blood-based breast cancer detection test based on functional enrichment of breast adenocarcinoma associated circulating tumor cells (BrAD-CTCs) and their identification via multiplexed fluorescence immunocytochemistry (ICC) profiling for GCDFP15, GATA3, EpCAM, PanCK and CD45 status.

**RESULTS:** The test accurately detects BrAD-CTCs in breast cancers irrespective of age, ethnicity, disease stage, grade and hormone receptor status. Analytical validation established the high accuracy and reliability of the test under intended use conditions. A case-control study with samples from 9,632 healthy women and 548 known BrC cases established 100% specificity and 92.07% overall sensitivity; stage-wise sensitivities were 70.00% for Stage 0, 89.36% for Stage I, 95.74% for Stage II and 100% for Stage III and Stage IV cancers. In a prospective clinical study with 141 suspected cases of breast cancer who underwent a biopsy after blood collection, the test showed 93.1% specificity and 94.64% overall sensitivity in differentiating breast cancer cases (n = 112) from those with benign breast conditions (n = 29); stage-wise sensitivities were 87.50% for Stage 0, 95.83% for Stage I and Stage II, 95.00% for Stage III and 100% for Stage IV cancers.

**CONCLUSION:** The findings reported in this manuscript support the clinical potential of this test for blood based BrC detection.

## BACKGROUND

Breast Cancer (BrC) is the most common malignancy and a leading cause of cancer related mortality among women globally (1). Although mammography is the standard of BrC screening in asymptomatic females, there is a need for improved BrC detection which addresses the risks and limitations of mammography, such as radiation exposure, lower specificity in differentiating benign conditions from malignancy, lower sensitivity for invasive carcinomas, as well as incompatibility with dense breast tissue. Circulating tumor analytes in peripheral blood have been evaluated for potential application in more accurate, non-radiological and non-/ minimally invasive screening for breast cancer. Circulating tumor cells (CTCs) are an ideal analyte for detection of cancers since they are intact malignant cells which harbour the imprint of the parent tumor. CTCs have distinct advantages over nucleic acid fragments or serum antigens since the latter may also be released by non-malignant cells and are associated with lower sensitivity and specificity respectively. There is evidence of sufficient viable CTCs being released into blood even during early stages of carcinogenesis. In breast cancer, it is reported that angiogenesis commences at the DCIS stage itself which can facilitate the dissemination of tumor cells (2–4). Such disseminated tumor cells (DTCs) have been reported in bone marrow of 20% to >50% of patients with DCIS or DCIS with microinvasion, respectively (5–7). Prior studies have also indicated high detection rates of CTCs in blood samples of patients with early-stage breast cancers. Using nanostructured coated slides, Krol et al (8) reported 62.5% CTC detection rate for Stage I and II BrC. Using filtration-based devices, Reduzzi et al (9) showed 76% CTC detection rate in early-stage breast cancer. Similarly, Jin et al (10) used the CytoSorter^®^ CTC capture system and showed 50% and >80% sensitivity in DCIS and Stage I / II BrC. These studies support the biological plausibility of CTC-based cancer screening approaches. Although CTCs have been evaluated for cancer detection, the inability of prior technologies to effectively enrich and harvest sufficient CTCs has hindered meaningful downstream applications. Most prior attempts at evaluating CTCs for cancer screening have been based on epitope capture using the CellSearch platform which, while not approved for CTC detection, finds frequent research use. Several prior studies have highlighted the lower performance of epitope capture arising due to its inability to efficiently harvest or detect CTCs with lower expression of EpCAM and PanCK which are the most routinely employed target markers (11–17). We have previously described a novel functional enrichment method with high CTC detection sensitivity, which yields sufficient CTCs for downstream applications such as immunocytochemistry (ICC) profiling (18,19). In this manuscript we describe the validation of this technology for use as BrC detection test. We show that the technology has vastly improved CTC detection sensitivity even in stage 0 BrC (DCIS) and addresses several limitations of prior CTC based cancer detection efforts.

## METHODS

### Study Participants and Samples

Samples for method development and validation were obtained from participants in two ongoing observational studies of the sponsor, TRUEBLOOD (WHO ICTRP https://trialsearch.who.int/?TrialID=CTRI/2019/03/017918, CTRI http://ctri.nic.in/Clinicaltrials/pmaindet2.php?trialid=31879) and RESOLUTE (WHO ICTRP https://trialsearch.who.int/?TrialID=CTRI/2019/01/017219, CTRI http://ctri.nic.in/Clinicaltrials/pmaindet2.php?trialid=30733) the design of which were intended to support the identification and characterization of blood-based malignant tumor derived analytes for non-/ minimally invasive cancer detection. The TRUEBLOOD study enrolled known cases of cancers and symptomatic individuals suspected of solid organ cancers. The RESOLUTE study enrolled asymptomatic adults with no prior diagnosis of cancer, no current symptoms or findings suspected of cancer and only age associated risk of cancer. Both studies were approved by the Ethics Committees of the sponsor (Datar Cancer Genetics, DCG) as well as the participating institutes and were performed in accordance with the Declaration of Helsinki. Fifteen millilitres of peripheral blood were collected from all enrolled study participants in EDTA vacutainers after obtaining written informed consent. Where possible, tissue samples were also obtained from TRUEBLOOD study participants posted for a biopsy as per standard of care (SoC) procedures (tissue samples were used for method development). In addition, leftover blood samples from suspected or known (recently diagnosed or pre-treated) cancer patients who availed of the study sponsor’s commercial services for cancer management as well as healthy (asymptomatic) volunteers at the study sponsor’s organization were also obtained after due consent. Blood samples (15 mL) from suspected cases of cancers were collected prior to the patients undergoing an invasive biopsy. All biological samples were assigned alphanumeric barcodes and stored at 2°C −8°C during transport to reach the clinical laboratory within 46 h. Sample blinding avoided systematic differences between groups due to (un)known baseline variables that could affect the test findings and also eliminated potential biases that could have otherwise arisen due to operator’s knowledge of the sample. From the originally collected 15 mL blood samples, a 5 mL aliquot was set aside for processing (CTC enrichment and ICC profiling) as part of clinical studies. The remaining blood samples were used for various method development studies. All samples were processed at the CAP and CLIA accredited facilities of the Study Sponsor Datar Cancer Genetics, which also adhere to quality standards ISO 9001:2015, ISO 27001:2013 and ISO 15189:2012.

### Enrichment of Circulating Tumor Cells from Peripheral Blood

Aliquoted blood samples (5 mL) were processed for the enrichment of CTCs from peripheral blood mononuclear cells (PBMC) as described previously (20). Comprehensive details are provided in Supplementary Materials.

### Immunocytochemistry Profiling of CTCs

The process of ICC profiling of CTCs was as described previously (19). Comprehensive details are provided in Supplementary Materials. Figure 1 is a Schema of the test showing the various steps in CTC enrichment and identification by ICC profiling for various markers. The Decision Matrix for assigning samples as Positive, Equivocal or Negative based on the findings of ICC profiling is provided in Figure 2.

**Figure 1.**
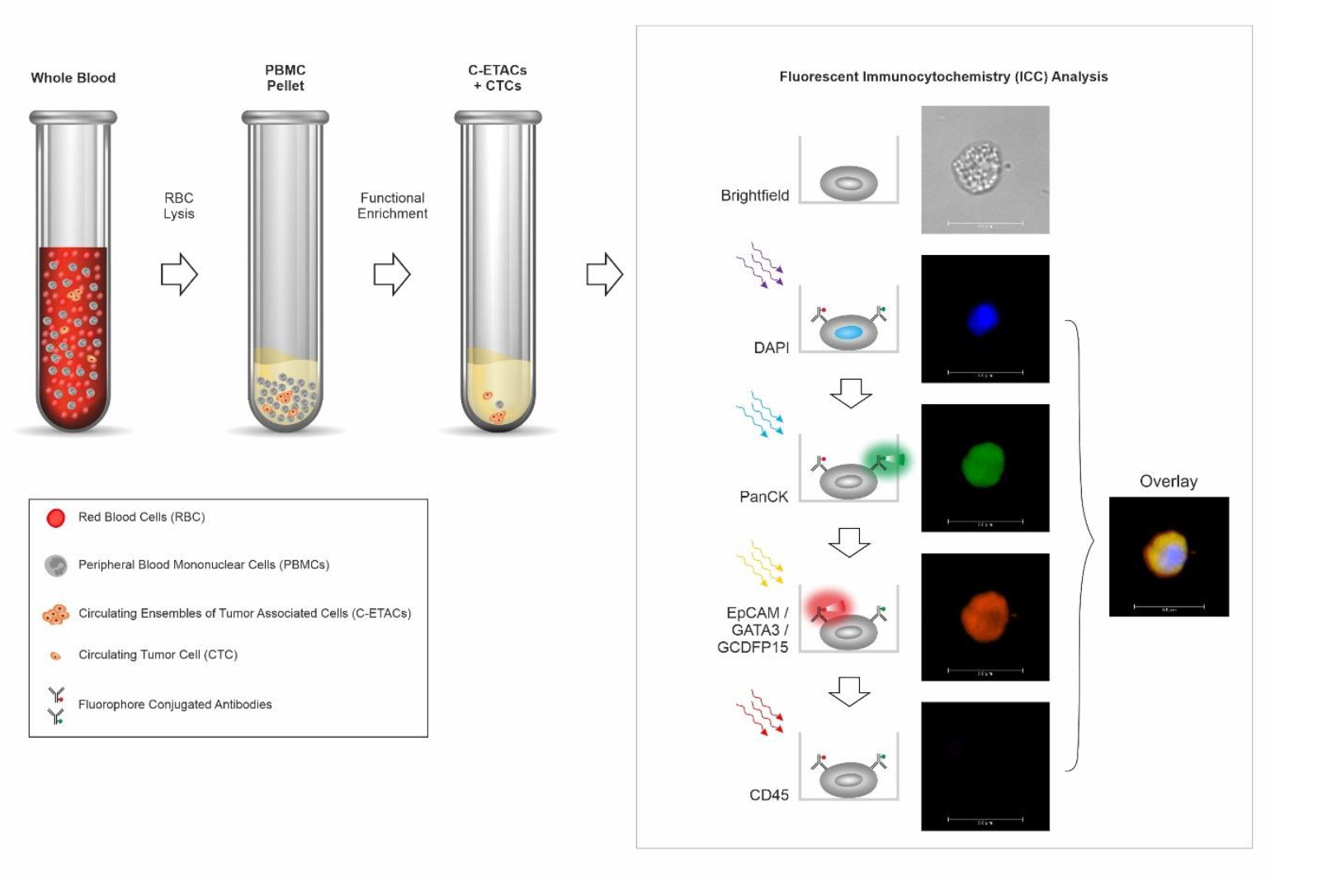
Schema of Test. Functional enrichment of circulating tumor cells (CTCs) is achieved using a cell culture medium that is cytotoxic towards all non-malignant cells and permits survival of tumor derived malignant cells. Peripheral blood mononuclear cells (PBMC) isolated from whole blood are treated with the custom proprietary medium for 120 h after which the surviving cells and cell clusters are harvested and evaluated by multiplexed immunocytochemistry (ICC) profiling to determine presence of breast adenocarcinoma associated CTCs (BrAD-CTCs) which are identified as CD45-negative cells which express GATA3, GCDFP15 and EpCAM in combination with PanCK.

**Figure 2.**
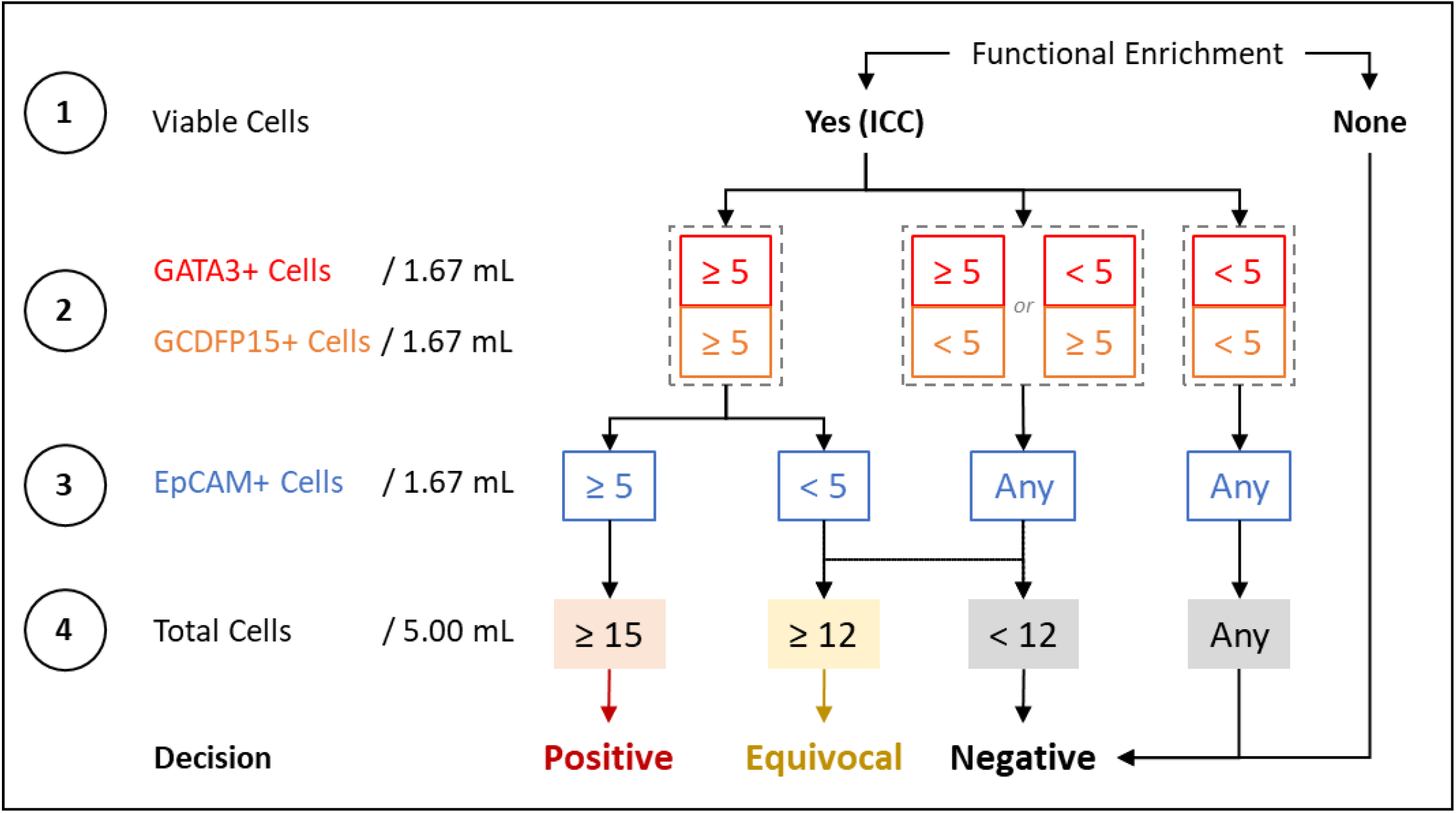
Decision Matrix for Classifying Samples. The detection threshold for breast adenocarcinoma associated CTCs (BrAD-CTCs) is ≥ 15 PanCK cells / 5 mL, which is constituted by the detection of ≥ 5 GATA3+, PanCK+, CD45-cells along with ≥ 5 GCDFP15+, PanCK+, CD45-cells as well as ≥ 5 EpCAM+, PanCK+, CD45-cells in the respective aliquots. Depending on the number of each type of marker positive cells, samples are marked as Positive, Equivocal or Negative. The decision matrix bestows priority to GATA3 and GCDFP15 over EpCAM while classifying samples to ensure specificity for BrAD over other epithelial malignancies where EpCAM+ cells may be detected but breast-specific markers would be absent. Thus, while the test can detect EpCAM+, PanCK+, CD45-cells which may be present in various epithelial malignancies, it specifically reports only BrAD-CTCs.

### Method Development and Optimization

Comprehensive details of method development and optimization studies are provided in the Supplementary Materials.

### Analytical Validation

Analytical validation established the performance characteristics of the test with standard analyte (SKBR3 cells), spiked into healthy donor blood to generate various dilutions (cell densities). These dilutions were processed as per the described procedures (proprietary differentially cytotoxic medium treatment and ICC profiling) to determine the yield of spiked cells. Comprehensive details of analytical validation studies are provided in the Supplementary Materials.

### Case Control Clinical Study

The ability of the Test to discern / identify BrC from asymptomatic individuals was initially ascertained and established in a case control study with 548 females who were recently diagnosed, therapy naïve cases of BrC and 9,632 healthy females with no prior diagnosis of any cancer, no current suspicion of any cancer and with BIRADS-I on a mammogram, i.e., no evidence of breast cancer (Supplementary Table S1). Initially, samples in the Asymptomatic cohort were randomized into Training and Test Sets in a 70%:30% ratio. The BrC cases were first segregated by Stage (0 – IV) and the samples per stage were then assigned to Training and Test Sets in a 70%:30% ratio. The Training Set samples (384 BrC and 6742 cancer-free females) was initially evaluated with the analysts unblinded to the status of the samples to determine the concordance between the clinical status and the interpretation of the marker status based on the Decision Matrix. Then the blinded Test Set comprising of 164 BrC and 2890 cancer-free females’ samples was evaluated to determine the performance characteristics. Subsequently all Training and Test samples (BrC and healthy) were shuffled and random 30% samples (with stage-wise for cancer) were selected for analysis as Test Set Iteration 2. This shuffling step was repeated to obtain 20 iterations of the Test Set. From these iterative 20 sets, median and range of sensitivity, specificity and accuracy were determined.

### Prospective Clinical Study

The performance characteristics of the test was next ascertained and established in a prospective blinded study of 141 individuals with clinical symptoms or radiological findings, who were referred for a biopsy due to suspicion of breast cancer (Supplementary Table S2). All participants provided blood sample prior to the biopsy. The sponsor was blinded to the diagnosis, i.e., the findings of the histopathological examination (HPE). Samples were prospectively accrued in this study until 24 samples each were obtained for Stage 0, I and II, 20 samples each were accrued for Stage III and IV and 29 samples were accrued for individuals with benign findings. Clinical status of samples (cancer / benign) was revealed to sponsor only after sample analysis was complete and test findings shared with the clinical study investigator. From these samples, performance characteristics including sensitivity, specificity and accuracy were determined with equivocal findings considered as positive and as negative, respectively.

### Molecular Concordance Study

In a combined subset of 61 samples from the case control and prospective cohorts, where matched tumor tissue and blood samples were available, a molecular concordance study was performed. Tumor Tissue DNA (ttDNA) was isolated and profiled by Next Generation Sequencing (NGS) using the Ion Proton Platform and the Comprehensive Ampliseq Multi (409)-gene Cancer Panel. Simultaneously, PBMCs were isolated from the matched blood samples and used for CTC enrichment. On the 5^th^ day, genomic DNA (gDNA) isolated from all surviving cells was evaluated by a ddPCR assay specific to the driver mutation on a BioRad QX200 platform. Concordance between tumor tissue and CTCs was determined as the proportion of the latter where the corresponding gene variant was detected by ddPCR.

## RESULTS

### Method Development and Optimization

The method development and optimization studies showed the viability of multiplexed fluorescence analysis of markers with minimal or no cross-interference of markers as well as the ability to detect CTCs with much lower marker expression than primary tumor cells or reference cell lines. Additionally the studies also showed the capability of the test in detecting CTCs irrespective of patient age, ethnicity, cancer stage, tumor grade, subtype and hormone receptor status. Findings of method development and optimization are provided in the Supplementary Materials.

### Analytical Validation

The analytical validation studies established the analyte stability and also demonstrated the high sensitivity and specificity of the test as well as significant linear characteristics in addition to high precision. The sensitivity of the test was not adversely affected in presence of potentially interfering substances or by controlled variations to operating parameters. Findings of analytical validation which established these performance characteristics of the test are provided in the Supplementary Materials. The summary of the analytical validation studies is provided in Table 1.

**Table 1.** Summary of Analytical Validation Studies. The summary of findings of the Analytical Validation Studies indicate that the Test provides consistent, accurate and reproducible results with little or no interference from routine endogenous or exogenous factors when samples are obtained, stored and processed under the recommended conditions.

|  | EpCAM,<br>PanCK, CD45 | GATA3,<br>PanCK, CD45 | GCDFP15,<br>PanCK, CD45 | Overall |
| --- | --- | --- | --- | --- |
| Analyte Stability | 48 h |  |  |  |
| Recovery<sup>1</sup> | 94.6% | 86.4% | 88.6% | 89.9% |
| Limit of detection | 1 cell / mL |  |  |  |
| Linear Range | 1 - 64 cells / mL |  |  |  |
| Linearity | R<sup>2</sup> ≥ 0.98 | R<sup>2</sup> ≥ 0.98 | R<sup>2</sup> ≥ 0.98 | R<sup>2</sup> ≥ 0.98 |
| Sensitivity | 96.0%<br>(86.3% - 99.5%) | 98.0%<br>(89.4% - 99.9%) | 94.0%<br>(83.5% - 98.8%) | 94.0%<br>(83.5% - 98.8%) |
| Specificity | 100.0%<br>(88.4% - 100.0%) | 100.0%<br>(88.4% - 100.0%) | 100.0%<br>(88.4% - 100.0%) | 100.0%<br>(88.4% - 100.0%) |
| Accuracy | 97.5%<br>(91.3% to 99.7%) | 98.8%<br>(93.2% to 99.9%) | 96.3%<br>(89.4% - 99.2%) | 96.3%<br>(89.4% - 99.2%) |
| Precision | CV = 4.6% | CV = 3.9% | CV = 3.8% | CV = 4.1% |
| Robustness | >95% |  |  |  |
| <sup>1</sup>Above 10 cells / 5 mL as determined from the Linearity experiment. Values within parentheses represent 95% CI. |  |  |  |  |

### Case Control Clinical Study

We evaluated the performance characteristics of the test in two clinical studies. In the clinical study which was a case control cross-validation study, the median stage wise sensitivities were as follows: 70% for Stage 0, 89.36% for Stage I, 95.74% for Stage II, 100% for Stage III,100% for Stage IV and 92.07% overall. In absence of any positive of equivocal findings in the control (cancer free and asymptomatic) cohort, the specificity of the test (cancer v/s healthy) was 100%. Cancer samples (cases) with equivocal findings were considered as positive for determination of sensitivity and accuracy. Table 2 provides the specificity as well as median of stage-wise and cumulative sensitivity and accuracy across the 20 iterations. Details of this iteration analysis are provided in Supplementary Table S3. Sensitivity and accuracy were also determined with samples with equivocal findings being considered as negative. These findings are presented in Supplementary Table S4, which also indicates the stage-wise and cumulative range of sensitivity and accuracy.

**Table 2.** Summary of Clinical Validation Studies. The table provides the summary of both clinical validation studies. The stringent cross validation design of the case control (cancer v/s healthy) study yielded a range of sensitivities and accuracies, the median of which are reported along with the 95% confidence interval (CI) for the median. Cancer samples (cases) with equivocal findings were considered as positive for determination of sensitivity and accuracy. The prospective clinical study evaluated the performance of the test among a cohort of symptomatic cases who were eventually diagnosed with breast cancer or benign conditions of the breast. In this study, benign samples with equivocal findings were considered as false positives for determination of specificity and accuracy. Additional analyses are provided in Supplementary Tables S3 – S6.

|  | **Case Control Study, Cancer v/s Asymptomatic**<br>*Specificity: 100.00% (95% CI: 99.87% - 100.00%)* |  | **Prospective Study, Cancer v/s Benign**<br>*Specificity: 93.10% (95% CI: 77.23% - 99.15%)* |  |
| --- | --- | --- | --- | --- |
|  | **Sensitivity** | **Accuracy** | **Sensitivity** | **Accuracy** |
| **Cumulative** | 92.07%<br>95%CI: 91.12% - 93.03% | 99.57%<br>95%CI: 99.34% - 99.81% | 94.64%<br>95%CI: 88.70% - 98.01% | 94.33%<br>95%CI: 89.13% - 97.52% |
| *Stage 0* | 70.00%<br>95%CI: 34.75% - 93.33% | 99.90%<br>95%CI: 99.70% - 99.98% | 87.50%<br>95%CI: 67.64% - 97.34% | 90.57%<br>95%CI: 79.34% - 96.87% |
| *Stage I* | 89.36%<br>95%CI: 76.90% - 96.45% | 99.81%<br>95%CI: 99.60% - 99.94% | 95.83%<br>95%CI: 78.88% - 99.89% | 94.34%<br>95%CI: 84.34% - 98.82% |
| *Stage II* | 95.74%<br>95%CI: 85.46% - 99.48% | 99.91%<br>95%CI: 99.75% - 99.99% | 95.83%<br>95%CI: 78.88% - 99.89% | 94.34%<br>95%CI: 84.34% - 98.82% |
| *Stage III* | 100.0%<br>95%CI: 88.43% - 100.00% | 100.0%<br>95%CI: 99.87% - 100.00% | 95.00%<br>95%CI: 75.13% - 99.87% | 93.88%<br>95%CI: 83.13% - 98.72% |
| *Stage IV* | 100.0%<br>95%CI: 88.43% - 100.00% | 100.0%<br>95%CI: 99.87% - 100.00% | 100.00%<br>95%CI: 83.16% - 100.00% | 95.92%<br>95%CI: 86.02% - 99.50% |

### Prospective Clinical Study

The second study was an independently conducted blinded prospective study. Of the total 141 individuals from whom samples were collected, there were 112 breast cancer cases (Stages 0 – IV) and 29 cases of various benign breast conditions. There were no samples with equivocal findings in the cancer cohort hence the overall sensitivity was 94.6% with stage wise sensitivities being 87.5% for Stage 0, 95.8% for Stage I, 95.8% for Stage II, 95.0% for Stage III and 100% for Stage IV. Two samples with equivocal findings were diagnosed with benign conditions of the breast. In absence of follow-up data indicating if these cases were indeed subsequently diagnosed with breast cancer, the samples were considered as false positives (worst case scenario), based on which the specificity of the test (cancer v/s benign) was determined to be 93.1%. When samples with equivocal findings were considered as negative, the specificity of the test (cancer v/s benign) was 100% (best case scenario). The sample-wise details of the prospective validation cohort findings are provided in Supplementary Table S5. The stage wise and cumulative sensitivity and accuracy for both these scenarios are provided in Supplementary Table S6.

### Molecular Concordance Study

We identified a subset of 61 samples where driver mutations (allele frequency > 0.14) were detected by NGS in tumor tissue; for variants detected in 53 samples a specific TaqMan ddPCR assay was available. CTC-enriched fraction from these samples was used for gDNA isolation which in turn was evaluated by a ddPCR assay specific to the driver mutation on a BioRad QX200 platform. Variants in ttDNA detected by NGS were also detected by ddPCR in 81.1% of CTCs indicating significant concordance (Supplementary Table S7).

## DISCUSSION

We describe a blood test for BrC detection in asymptomatic women based on multiplexed fluorescence ICC profiling of CTCs in peripheral blood. The test can accurately determine the presence of CTCs in BrC irrespective of stage, grade, subtype, age, ethnicity or hormone receptor status (Supplementary materials). Analytical validation established high sensitivity, specificity, precision and robustness in addition to non-interference from endogenous and exogenous factors (Supplementary materials). Two separate clinical studies established 100% specificity (cancer v/s asymptomatic) with 92% - 94% overall sensitivity and 70% - 87% Stage 0 sensitivity (Table 2). The test can differentiate samples from cancer patients and healthy individuals with high (100%) specificity and can also identify individuals with benign conditions with ≥93% specificity. Our test has (a) high sensitivity especially for early stages including DCIS, so that the majority of cancers are detected at localized stages which are amenable to curative resection, and (b) high specificity so that vast majority of cancer-free individuals do not undergo additional unnecessary procedures. Our test offers compelling advantages over screening mammography and is hence a strong candidate for non-invasive BrC screening in asymptomatic women.

Presently, any benefits of standard mammography screening are largely in populations aged 50 years and above who have a higher age associated cancer risk (21–27). Standard 2D digital mammography has been reported to have 73%-87.3% sensitivity and 86%-96% specificity (28–30). The low accuracy of screening mammography has been noted in younger women, particularly those aged below 40 years (31). In addition, challenges associated with screening mammography are the high rates of false positives (7-12% at first mammogram (32) and 50-60% after ten yearly mammograms (33)). Besides, mammography also has a lower sensitivity for invasive cancers (76%-85%) than DCIS (83.0%-94.3%) (29,30). Prior studies have also suggested a modest association between radiation exposure in mammograms and elevated risk of cancer in BRCA1 mutation carriers (34).

A longitudinal study of screening mammograms in 69,025 women reported 705 cases of screen detected BrC (SBC) and 206 cases of Interval BrC (IBC, not detected by mammography) (35); the latter were more likely to be of high grade as well as have higher mortality than SBC. Niraula et al encourage a re-evaluation of the concept of population-based screening mammography and recommend exploring strategies beyond conventional screening mammography. In support, the 2018 data available at the US National Centre for Health Statistics, Centres for Disease Control and Prevention indicates that approximately 27.1% of women in the age group 50 to 74 years defaulted on SoC mammography (36). Improved BrC screening and risk-mitigation strategies are hence vital to improve compliance and BrC detection.

Recent efforts at developing non-invasive cancer screening technologies have focussed on a multi-or a pan-cancer approach. Notably, GRAIL’s Galleri has introduced the pan-cancer screening test based on methylation profiling in ctDNA (37). However, the Galleri test has very low sensitivity (<10% - 16%) for Stage I BrC (38,39), with no data on its ability to detect DCIS. Similarly, the CancerSEEK test based on simultaneous evaluation of serum proteins and gene variants has ∼40% cumulative sensitivity for early stage and overall ∼30% sensitivity for BrC (40). Purposeful screening for early cancer detection necessitates sensitivity for early stages (0 – II), which has not been demonstrated by these tests. To the best of our knowledge, there are no other platforms with high sensitivity and specificity for early stages (0 – II) of BrC.

The present test is based on detection of CTCs, which are ubiquitously found in the blood of patients with an underlying solid organ cancer and are undetectable in healthy individuals (18,19). We show that functionally enriched CTCs can differentiate malignant conditions from benign conditions as well as identify asymptomatic individuals with an underlying malignant condition with high specificity. Obtaining numerically sufficient CTCs is akin to a non-invasive biopsy of the breast tumor without stromal or other non-tumor content. Since the present test is based on detection of CTCs which represent the hematogenous phase of carcinomas associated with a higher risk of progression / metastasis, it is likely that the test has a higher sensitivity for detecting those sub-populations of DCIS where the risk of progression is higher.

Traditionally, epitope capture with Anti-EpCAM has been the preferred method for CTC enrichment. However, several studies have demonstrated the poor CTC capture / detection rate of this platform (13,14,41). It would be pertinent to mention that although technologies like CellSearch find frequent mention in research pertaining to cancer detection, these have not been approved for detection of cancers based on CTCs. Hence the limitations of these technologies for cancer detection must be critically understood and proactively addressed to improve CTC and cancer detection; our test has been developed on such a working hypothesis. The label- and size-agnostic functional CTC enrichment technique in our test is immune to the limitations of epitope capture platforms and hence may offer a more realistic CTC detection rate. In our test, marker expression is determined by a sensitive high content screening (HCS) system with standardized thresholds to minimize false negatives. The detection thresholds of the test accommodate CTCs with significantly lower marker expression (as compared to tumor cells or reference cell lines), such as those undergoing epithelial to mesenchymal transition (42,43).

The potential benefits of the test include early BrC detection, especially in asymptomatic women who decline guideline recommended screening mammography as well as in asymptomatic women for whom the guidelines may not recommend routine screening mammography (e.g., those below 50 years of age). The high (>90%) cumulative sensitivity at Stage 0-II indicates a <10% risk of missing these localized cases where the disease has not spread to other organs and where the 5-year survival rate is ∼99%. For the <10% cases that are not detected at local stages, subsequent detection at Stage III (regional spread) with >95% reported sensitivity is still associated with ∼86% 5-year survival. The test also has a significantly higher sensitivity for invasive carcinomas (all stages) than has been reported for screening mammography (29,30), and can potentially mitigate risks of IBC (this is yet to be prospectively established). The 100% specificity of the test indicates a negligible risk of false positives. There are limited or no risks associated with use of the test since it is non-invasive and is performed on a venous blood draw of 5 mL of peripheral blood.

The strength of our study stems from the use of an adequately powered sample size and avoidance of overfitting, since the findings of the iterative validation study agreed well with that of the training set.

The test has certain limitations in the context of a universal BrC screening. The sensitivity of the Test was lowest for Stage 0 disease. However, this does not present any increased risk of false negatives as compared to screening mammography. Since individuals with potentially false negative findings would not be deprived of standard mammography screening, it would not add to the pre-existing risk of the individual. While there is virtually no risk of false positives, detection of BrAD-CTCs may be construed as false positive in individuals where the malignancy may not be immediately evident on a standard screening mammogram or in a biopsy (as was observed in the prospective validation cohort). This risk may be mitigated by use of a more diagnostically relevant imaging modality or follow-up among individuals with positive test findings. Minor non-(Adeno) carcinoma subtypes of breast cancers are not detected by this test. The test has not been evaluated in a prospective large cohort study in the intent to test asymptomatic population. Finally, as inherent to any cancer screening test, our test could result in over-diagnosis and over-treatment.

### CONCLUSION

We describe a blood-based, non-invasive test which detects breast AD associated CTCs with high specificity and sensitivity. The test presents a superior alternative to mammography screening of asymptomatic women for BrC detection. Approximately 38 million mammograms are performed every year in the US (44) of whom ∼280,000 (∼0.75%) of cases are diagnosed with BrC (1,45,46). Similarly, of the ∼16 million mammograms performed annually in Europe (47), ∼500,000 (1) (∼3.1%) are diagnosed with BrC. Our test has the potential to minimize the need for mammography screening in individuals with positive findings who could be referred for diagnostic imaging and work up. The test may also minimize the need for screening mammography in individuals with negative findings.

## Data Availability

All relevant data are included in the manuscript and its Supplementary Information file.

## ADDITIONAL INFORMATION

## ACKNOWLEDGEMENTS

The authors are grateful to the staff of the Study Sponsor (DCG) for their contributions in managing various clinical, operational and laboratory aspects of the study.

## AUTHOR CONTRIBUTIONS

Timothy Crook: Conceptualization, Investigation, Methodology, Resources, Supervision, Writing - review & editing; Darshana Patil: Conceptualization, Formal analysis, Investigation, Methodology, Project administration, Resources, Software, Supervision, Validation, Visualization, Writing - review & editing; Dadasaheb Akolkar: Conceptualization, Formal analysis, Investigation, Methodology, Project administration, Resources, Software, Supervision, Validation, Visualization, Writing - review & editing; Anantbhushan Ranade: Conceptualization, Methodology, Writing - review & editing; Sewanti Limaye: Conceptualization, Methodology, Writing - review & editing; Raymond Page: Conceptualization, Methodology, Writing - review & editing; Vineet Datta: Conceptualization, Investigation, Methodology, Project administration, Resources, Supervision, Writing - review & editing; Pradeep Fulmali: Conceptualization, Formal analysis, Methodology, Project administration, Resources, Supervision, Validation, Visualization, Writing - review & editing; Sachin Apurwa: Data curation, Formal analysis, Methodology, Resources, Software, Validation, Visualization, Writing - review & editing; Stefan Schuster: Conceptualization, Methodology, Writing - review & editing; Ajay Srinivasan: Conceptualization, Data curation, Formal analysis, Methodology, Software, Supervision, Validation, Visualization, Writing - original draft, Writing - review & editing; Rajan Datar: Conceptualization, Funding acquisition, Investigation, Methodology, Resources, Software, Supervision, Validation, Visualization, Writing - original draft, Writing - review & editing

### ETHICS APPROVAL AND CONSENT TO PARTICIPATE

All biological samples were obtained from participants in two studies, TRUEBLOOD (WHO ICTRP https://trialsearch.who.int/?TrialID=CTRI/2019/03/017918, CTRI http://ctri.nic.in/Clinicaltrials/pmaindet2.php?trialid=31879) and RESOLUTE (WHO ICTRP https://trialsearch.who.int/?TrialID=CTRI/2019/01/017219, CTRI http://ctri.nic.in/Clinicaltrials/pmaindet2.php?trialid=30733). Both studies were approved by Datar Cancer Genetics Limited Institutional Ethics Committee. All participants provided written informed consent. Both studies were performed in accordance with the Declaration of Helsinki.

### CONSENT FOR PUBLICATION

All study participants consented for publication of de-identified biological data. The present manuscript does not contain any personal or identifiable information or data of any participant.

### DATA AVAILABILITY

All relevant data are included in the manuscript and its Supplementary Information file.

### FUNDING

No external funding was obtained for this study. The entire study was funded by the Study Sponsor (DCG).

