## Supplementary file for "Accurate Screening for Early-Stage Breast Cancer by Detection and Profiling of Circulating Tumor Cells"

**SUPPLEMENTARY MATERIALS, METHODS AND FINDINGS**

**Markers**

The test identifies breast adenocarcinoma associated circulating tumor cells (BrAD-CTCs) as malignant apoptosis resistant cells positive for EpCAM, PanCK, GATA3 and GCDFP-15 and negative for CD45. Pan-Cytokeratins (PanCK) are a family of cytoplasmic structural proteins that are present in epithelial tumors and in CTCs. PanCK positivity along with absence of CD45 expression is the primary determinant of CTCs. Epithelial Cell Adhesion Molecule (EpCAM) are membrane antigens present on epithelial cells (and carcinomas) that function in cell adhesion. GATA Binding Protein 3 (GATA3) is a transcription factor (nuclear marker) which is commonly overexpressed in breast (up to 94%), salivary duct, skin and urothelial carcinomas, and less commonly in carcinomas of the lungs, liver, pancreas, stomach, kidneys, thyroid, endometrium and ovary. Gross Cystic Disease Fluid Protein 15 (GCDFP15) expression is commonly reported in breast (up to 90%) and salivary duct carcinomas, and less commonly in carcinomas of the lungs and prostate. Common Leucocyte Antigen (CD45) serves to discern the CTCs from CD45 positive haematolymphoid cells. The combination of GATA3 and GCDFP15 ensures a high specificity of organ localisation. Classically, this combination of markers is used to identify a Breast primary in case of metastatic progression or recurrence. Neither marker is reported to be expressed in normal breast parenchyma or non-malignant conditions of breasts. Co-expression of GATA3 and GCDFP15 is reported in salivary duct carcinoma (SDC), a rare cancer type with significantly lower (1/200^th^) incidence than breast cancer.

**Antisera and Cell Lines**

The antisera used included recombinant human (RH) Anti CD326 IgG1-Vio 615 (Miltenyi Biotech), RH Anti-CK-IgG1-Vio 515 (Miltenyi Biotech), RH Anti-CD45-IgG1-APCVio 770 (Miltenyi Biotech), Mouse Anti-GATA3 IgG1/k (Dako), Mouse Anti-GCDFP15 IgG1/k (Dako) and Anti-Mouse Alexa Fluor 594 (Invitrogen). The reference cell lines include SKBR3 (breast cancer), MOLT-3 (leukemia) and SW982 (synovial sarcoma) all of which were procured from ATCC. The purity of reference cell lines was confirmed by Short Tandem Repeat (STR) Profiling and testing for Mycoplasma every 6 months.

**Isolation of Primary Tumor Derived Cells**

The isolation of primary tumor derived cells (TDCs) from an excised tumor (malignant / benign) has been described previously (1).

**Enrichment of Circulating Tumor Cells from Peripheral Blood**

Aliquoted blood samples (5 mL) were processed for the enrichment of circulating tumor cells (CTCs) from peripheral blood mononuclear cells (PBMC) as described previously (1). Briefly, PBMCs were isolated from whole blood via lysis of red blood cells (RBCs) followed by centrifugation. PBMCs resuspended in Phosphate Buffered Saline (PBS) were treated with a proprietary differentially cytotoxic medium that induces cell death in all apoptosis-competent non-malignant (hemato-lymphoid, epithelial and endothelial) cells, while malignant tumor derived cells (CTCs) survive due to apoptosis resistance. After treatment for 5 days at 37°C, surviving cells and cell clusters are harvested by centrifugation and resuspended in PBS.

**Immunocytochemistry Profiling of Circulating Tumor Cells**

The process of ICC profiling of CTCs has been described previously (2). Briefly, CTCs enriched from 5 mL of blood were resuspended in 1500 μL 1x Phosphate Buffered Saline (PBS) and 100 μL aliquots of enriched CTCs seeded into 15 wells. Cells in each well were equivalent to 333 μL blood sample. Cells were fixed with 4% Paraformaldehyde, permeabilized with 0.3% Triton-X 100 and treated with 3% BSA (blocking). Cells were immunostained with each of the 3 separate Primary (1°) Ab cocktails for multiplexed analysis of the following combination of markers, (a) Anti-PanCK (1:500), Anti-CD45 (1:500), Anti-EpCAM (1:500), (b) Anti-PanCK (1:500), Anti-CD45 (1:500), Anti-GATA3 (1:4), (c) Anti-PanCK (1:500), Anti-CD45 (1:500), Anti-GCDFP15 (1:2). Samples for GATA3 and GCDFP15 were incubated with secondary (2°) anti-mouse Ab (1:500). PBS washes followed each Ab incubation step. Each marker combination was evaluated in 5 wells (333 μL × 5 = 1.67 mL equivalent of blood). Finally, cells were treated with 4’,6-Diamidino-2-phenylindole dihydrochloride (DAPI) for nuclear staining. Control samples (SKBR3 for EpCAM, GATA3 and GCDFP15 and MOLT3 for CD45) were included in each run. Samples were evaluated on the CellInsight High Content Screening (HCS) Platform to determine the Fluorescence Intensity (FI) for each marker. Marker expression was determined by the sequential excitation and acquisition of fluorescence signal.

Cells isolated from a primary benign or malignant tumor were resuspended in phosphate buffered saline (PBS) solution and ICC profiled similarly as described above for CTCs.

**Method Development**

*Detection Thresholds*

SKBR3 cells, MOLT-3 cells, SW982 cells, BrAD-CTCs, malignant breast tumor derived cells (M-TDCs) and benign breast tumor derived cells (B-TDCs) were immunostained for all markers (20 replicates). FI was recorded for each marker to determine relative expression of each marker per cell type.

Finding: The FI of PanCK, GATA3, GCDFP15 and EpCAM were higher in SKBR3, BrC-TDC (M-TDC) and BrAD-CTCs than in MOLT-3, SW982 and benign breast tumor cells (B-TDC). The FI of CD45 was significantly higher in MOLT-3 than in the other cell types. FI of markers was lowest in SW982 (Supplementary Figure S1). Based on these findings, the FI threshold for positivity was assigned as 70,000 (relative fluorescence units, RFU) for PanCK and EpCAM and 50,000 RFU for GATA3 and GCDFP15; these apply as a lower threshold for SKBR3 (positive control (PC) for all 4 markers), BrC-TDC and BrAD-CTCs where expression is essential for positivity. These FI thresholds accommodate CTCs with lower marker expression than the M-TDC or cell lines, such as CTCs undergoing epithelial to mesenchymal transition (EMT). For CD45 40,000 RFU was set as the upper threshold in SKBR3, M-TDC and BrAD-CTCs where expression is not expected and as the lower threshold for MOLT-3 (PC for CD45, negative control (NC) for all other markers). Additionally, numerical thresholds were defined as the proportion of cells staining positively for each marker for the acceptance of positive control (PC; >60%) and negative control (NC; <1%).

*Marker Specificity*

We determined the specificity of the marker combination to BrC by evaluating their expression in various CTCs. FI for GATA3 and GCDFP15 were evaluated after immunostaining of CTCs from Cancers of the Cervix, Esophagus, Kidney, Lung, Ovary, Pancreas and Stomach.

Finding: As expected, expression of GATA3 and GCDFP15 was lower (FI < 50,000 U) in all non-BrC CTCs (Supplementary Figure S2).

*Marker Expression in Breast Cancer*

FI for GATA3, GCDFP15, EpCAM, PanCK and CD45 were evaluated in subsets of BrAD-CTCs stratified by Age-Group (n=249), Ductal v/s Lobular subtype (n=219), Grade (n=99), Hormone Receptor Status (n=159) and Stage (n=162). FI of all markers was also evaluated in BrAD-CTCs from a Caucasian population (n=65) and from a South Asian population (n=225) to determine if there are any variances due to ethnicity.

Finding: As can be seen in Supplementary Figures S3 – S7, there were no significant variations in FI of any marker due to any of these intrinsic factors. Similarly, expression of markers in CTCs from Caucasian samples were compatible with (not lower than) the FI thresholds which were established in CTCs from South Asian samples (Supplementary Figure S8).

*BrAD-CTCs in Benign or Inflammatory Breast Conditions*

To determine the specificity of the Test to discern BrC from non-malignant conditions of the Breast, we evaluated blood samples from 91 recently diagnosed cases of benign or inflammatory conditions of the breast (Supplementary Table S8). Samples were processed for CTC enrichment and ICC profiling as described above.

Finding: BrAD-CTCs were not detected in any cases indicating a specificity of 100% (cancer v/s benign).

**Analytical Validation**

Analytical validation established the performance characteristics of the test with standard analyte (SKBR3 cells), spiked into healthy donor blood to generate various dilutions (cell densities). These dilutions were processed as per the described procedures (proprietary differentially cytotoxic medium treatment and ICC profiling) to determine the yield of spiked cells. A summary of findings of the analytical validations is provided in Table 1.

*Stability and Recovery*

To determine the Analyte Stability, 27 × 5 mL aliquots of healthy donor blood were spiked with ~15 SKBR3 cells each (final = 3 cells / mL) and stored at 2°C - 8°C. Of the 27 aliquots, 9 aliquots each were used immediately or after 24h and 48h. Of the 9 aliquots evaluated at each time point, 3 aliquots were used to determine recovery of each cell type (multiplexed marker combinations). Additionally, 3 × 5 mL blood was collected from 5 known CTC+ cases of BrAD; one sample was processed immediately (0h), one after 24h at 2°C - 8°C and the third after 48h at 2°C - 8°C. Recoveries at 0h were normalized as 100% and recoveries at 24h and 48h were determined relative to the 0h recovery.

Finding: In the spiked samples, the recovery of EpCAM+ cells was 100.0%, 97.8% and 97.8% at 0h, 24h and 48h storage at 2°C - 8°C. Similarly, the recovery of GATA3+ cells was 100.0%, 93.3% and 91.1% at 0h, 24h and 48h, and that of GCDFP15+ cells was 100.0%, 100.0% and 97.8% at 0h, 24h and 48h (Supplementary Table S9). In clinical samples, the overall (combined PanCK+) recovery was 91.5% and 88.6% after 24h and 48h storage at 2°C - 8°C respectively when 0h recovery was normalized as 100% (Supplementary Table S10). The findings of the stability and recovery study indicated that the samples could be stored at 2°C-8°C for up to 48h with <15% loss of cells.

*Linearity*

SKBR3 cells were spiked into 264 × 5 mL aliquots of healthy donor blood samples, stored for 48h at 2°C - 8°C and then processed for recovery. The 264 aliquots comprised 3 sets of 88 aliquots (11 spikes × 8 replicates). The study also included 24 × 5 mL aliquots (3 sets × 8 replicates) of healthy donor blood samples which were not spiked. Each set was assigned to either of the 3 multiplexed marker combinations. Samples were stored for 48h at 2°C - 8°C. Linearity was evaluated by Linear Regression.

Finding: Recoveries of spiked cells were generally higher at spike densities of 5 cells / 5 mL and higher (Supplementary Figure S9). Coefficient of Determination (R^2^) ≥0.98 in all markers indicated a significant linear response, especially in the range of 5 - 1280 cells / 5 mL.

*Limits of Detection, Quantitation and Blank*

The Limit of Blank (LoB) was determined from the 24 × 5 mL unspiked healthy female donor blood samples in the Linearity study. The Limit of Detection (LoD) was determined from a subset of the Linearity Study which included 72 × 5 mL samples spiked with 1, 3 or 5 SKBR3 cells (24 each). The Limit of Quantitation (LoQ) was determined from a subset of the Linearity Study which included 96 × 5 mL samples spiked with 1, 3, 5 or 10 SKBR3 cells (24 each).

Finding: No GATA3+, GCDFP15+ or EpCAM+ cells were detected in the unspiked samples, i.e., no false positives. Thus, the limit of blank (LoB) was determined to be 0 cells / mL. The limit of detection (LoD) was 1 cell / 5 mL. The limit of quantitation (LoQ) was 10 cells / 5 mL, based on ≥80% recovery in at least 7 of 8 replicates.

*Sensitivity, Specificity and Accuracy*

SKBR3 cells were spiked into 50 × 15 mL aliquots (5 spikes × 10 replicates) of healthy donor blood at 15, 30, 60, 120 and 240 cells. Each 15 mL sample was split into 3 × 5 mL aliquots that were used for the analysis of each of the 3 marker combinations. Samples were stored for 48h at 2°C - 8°C prior to analysis. Unspiked healthy donor blood samples (30 × 5 mL) were included in this study for the determination of specificity. Samples with equivocal findings were considered as negative. Accuracy was determined based on total true positive and true negative samples detected out of the total 80 samples.

Finding: Among the 50 spiked samples evaluated for sensitivity, SKBR3 cells were detected in 47 samples, yielding a sensitivity of 94%. Since SKBR3 cells were undetectable in any of the 30 unspiked samples, the specificity was deemed to be 100%. (Supplementary Table S11). Accuracy, determined as the combined proportion of true positives and true negatives, was 96.3%.

*Precision*

On Day 1, User 1 spiked 15 (Low) SKBR3 cells into each of 8 × 5 mL aliquots of healthy donor blood, and 150 (High) SKBR3 cells into each of another 8 × 5 mL aliquots of healthy donor blood. All samples (8 Low spike + 8 High) were stored for 48h at 2°C - 8°C and processed. User 1 performed this study on 10 consecutive days and used one of two HCS Instruments. User 2 independently replicated the study on 10 consecutive days and used the second HCS Instrument. Mean Recoveries (%) were used to calculate Standard Deviation (SD) and Coefficient of Variation (CV, %) for Intra-Run, Inter-Run and Inter-Operator.

Finding: Precision of the test was determined across 2 operators in samples with high and low spike densities. Supplementary Table S12 provides the Coefficient of Variation (CV, %) for intra-run, inter-run and inter-operator precision for low and high spike as well as the cumulative for all markers. The cumulative CV was ≤4% for intra-run, ≤0.5% for inter-operator and ≤1.9% for inter-run precision. The overall CV was 3.8% indicating high precision.

*Robustness*

Guard-band studies were performed to ascertain and establish the robustness of the assay by varying operating parameters within predefined limits. Criteria evaluated include incubation temperatures, incubation times, centrifugation speeds, buffer volumes, and antibody dilutions (Supplementary Table S13). Around 15 SKBR3 cells were spiked into 5 mL healthy donor blood samples which were processed as per the Test procedure. Each Guard-band parameter was evaluated with 9 samples, where 3 samples each were evaluated at (a) the normal range, (b) the higher range and (c) the lower range. Each parameter passed the Guard-band test if the variance was <10%.

Finding: The overall variability ranged from 0.9% - 5.9% indicating that controlled changes to test parameters do not adversely impact the performance. During routine test conditions, these variations are controlled via the use of standard operating procedures (SOP) and instrument calibration.

*Interfering Substances*

The performance characteristics of the Test were evaluated in presence of endogenous (pathology markers) and exogenous factors (non-anticancer drugs) as potential interfering agents (Supplementary Table S14). Pure (analytical grade) molecules for each of these agents were obtained from commercial vendors and stored under recommended conditions until use. All substances were reconstituted as per manufacturer’s instructions in appropriate solvents to prepare working stock solutions which were immediately used for spiking studies. All exogenous substances (drugs) were used at the reported medically relevant Peak Plasma Concentrations (C_Max_) as per previously published literature, while endogenous substances (serum parameters) were evaluated at concentrations that are considered clinically elevated. Blood from a healthy donor (75 mL) who was not under any medication (last 14 days) was procured from a blood bank and spiked with about 750 SKBR3 cells. The spiked sample was split into 25 × 3 mL aliquots; 21 aliquots were spiked with each of the above substances at the indicated concentrations and 4 aliquots were used as unspiked controls. Each 3 mL sample was split into 3 × 1 mL aliquots; one aliquot each was used for detection of PanCK+, EpCAM+ cells; PanCK+, GATA3+ cells and PanCK+, GCDFP15+ cells respectively.

Finding: The presence of drugs at medically relevant peak plasma concentrations (C_Max_) or the deranged serum parameters did not impact the recovery or detection of SKBR3 cells spiked into blood samples.

**SUPPLEMENTARY FIGURES**

**Supplementary Figure S1. Detection Thresholds.**

SKBR3 (breast cancer) cells, MOLT3 (leukemia) cells, SW982 (sarcoma) cells, BrAD-CTCs, malignant breast tumor derived cells (M-TDCs) and benign breast tumor derived cells (B-TDCs) were immunostained to determine the expression level (FI: fluorescence intensity) of each marker.

**A. GATA3**

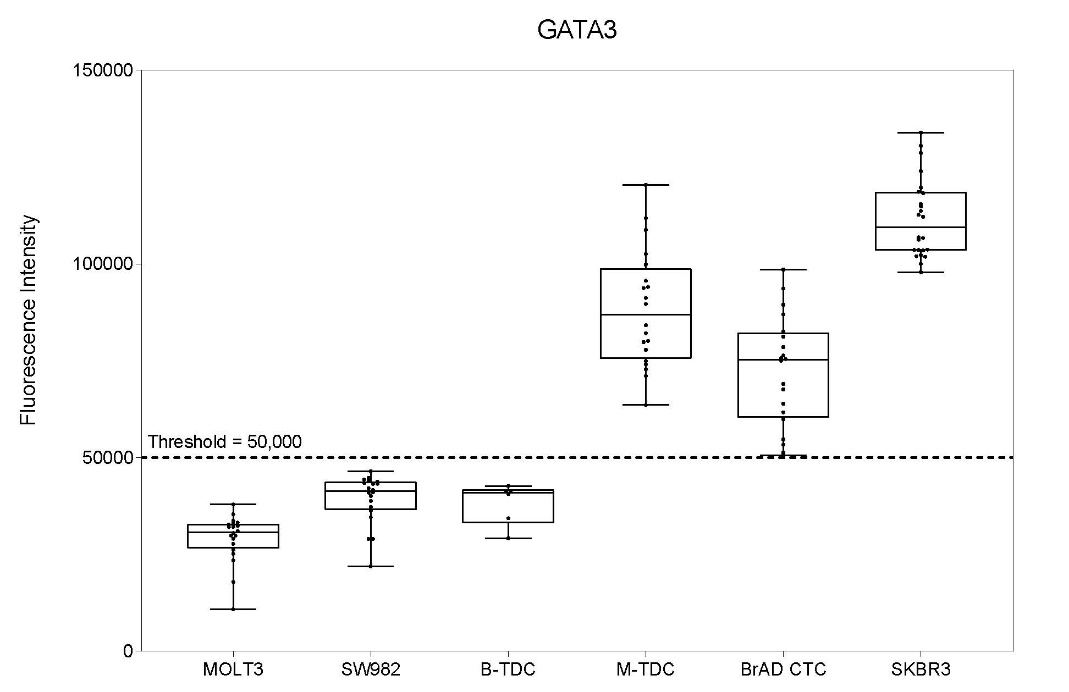

**B. GCDFP15**

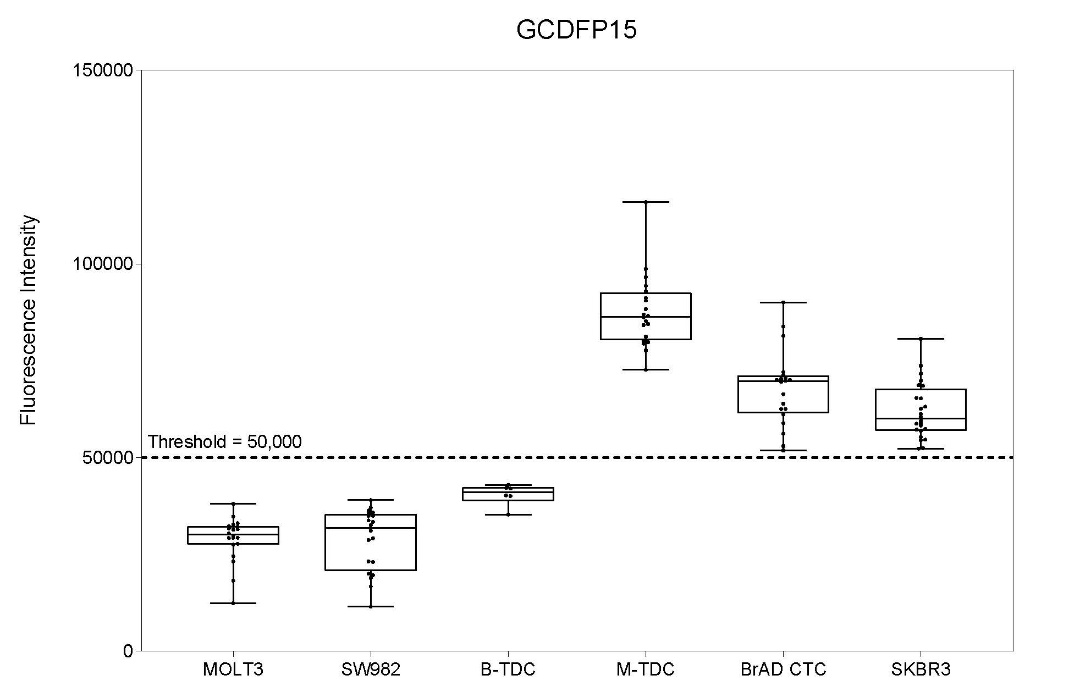

**C. EpCAM**

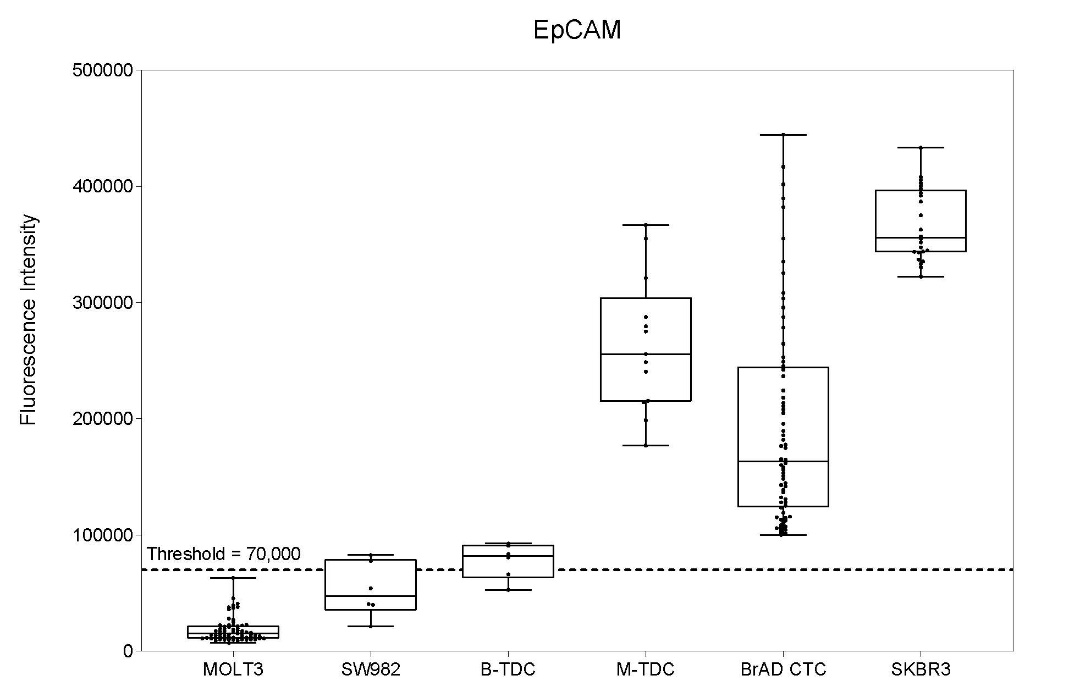

**D. PanCK**

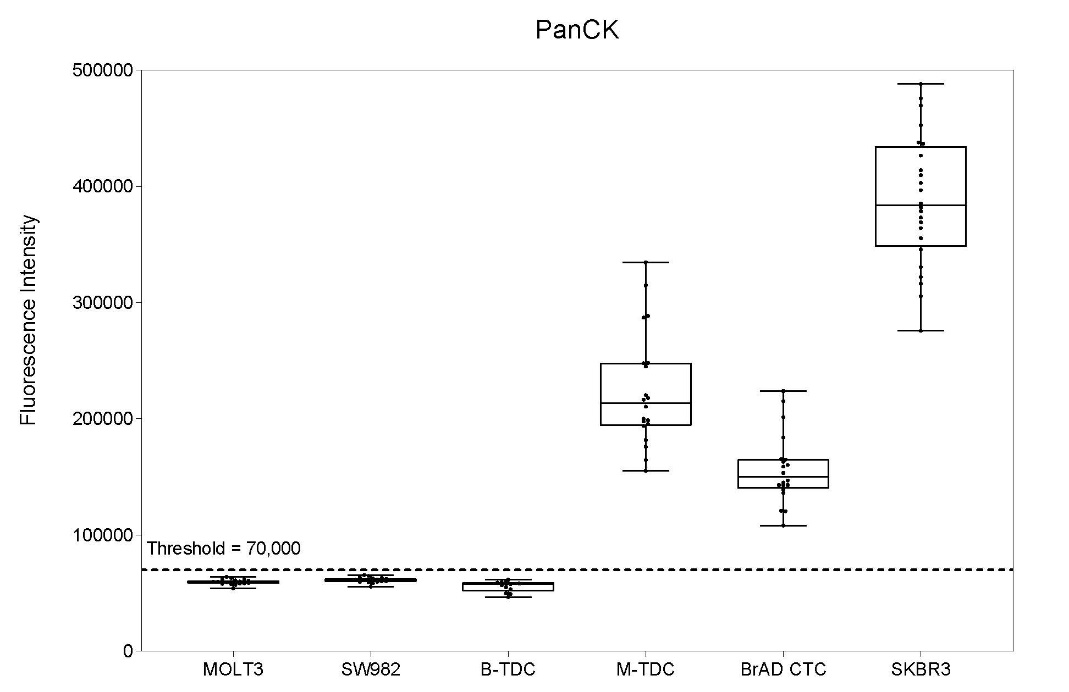

**E. CD45**

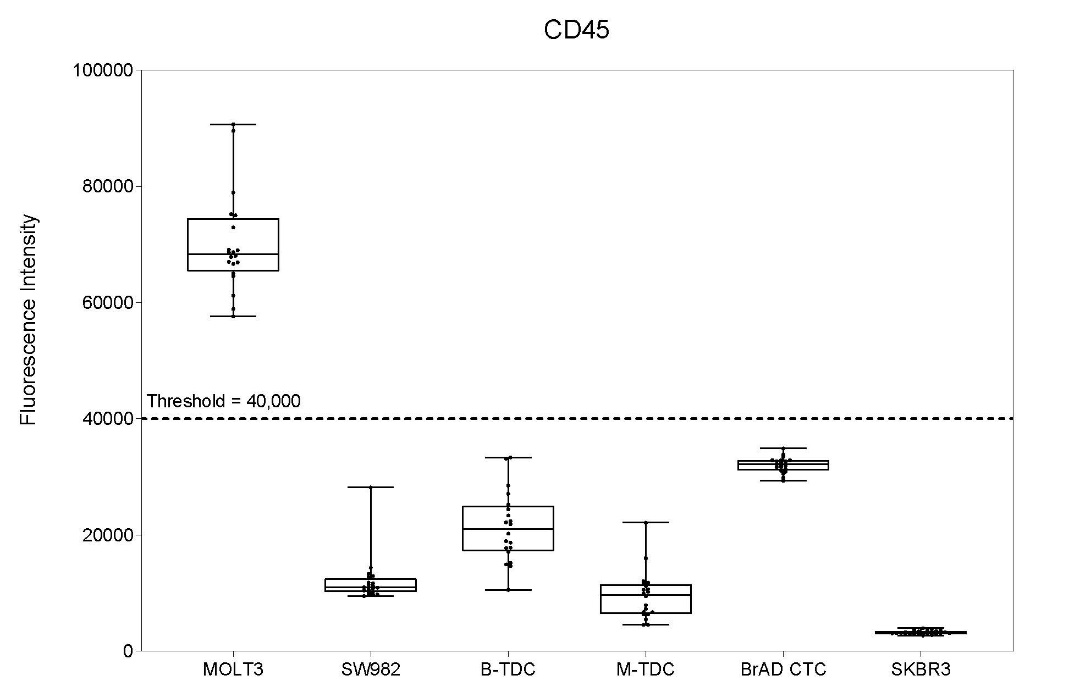

**Supplementary Figure S2. Expression of GATA3 (A) and GCDFP15 (B) in various CTCs.**

**A. GATA3 in various CTCs**

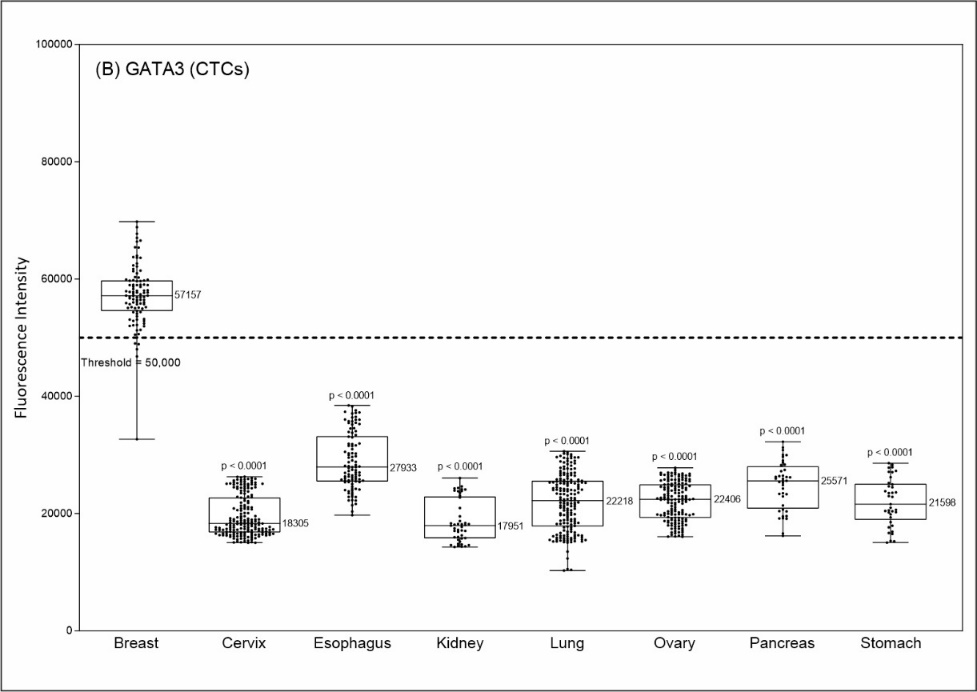

**B.** **GCDFP15 in various CTCs**

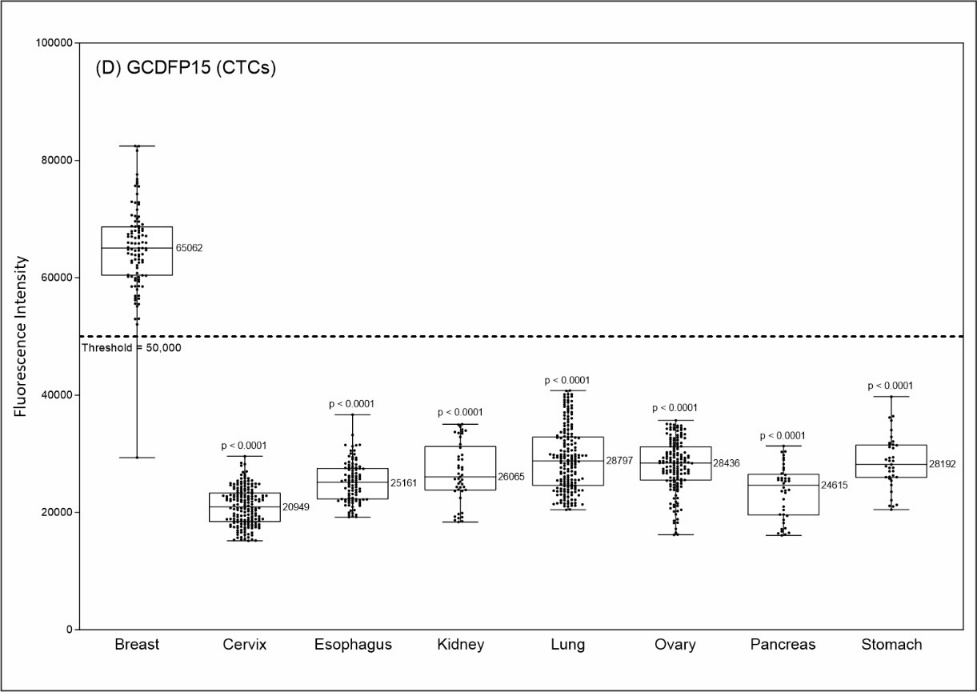

**Supplementary Figure S3. Age-group and Marker Expression on CTCs.** (A) GATA3, (B) GCDFP15, (C) EpCAM, (D) PanCK, (E) CD45

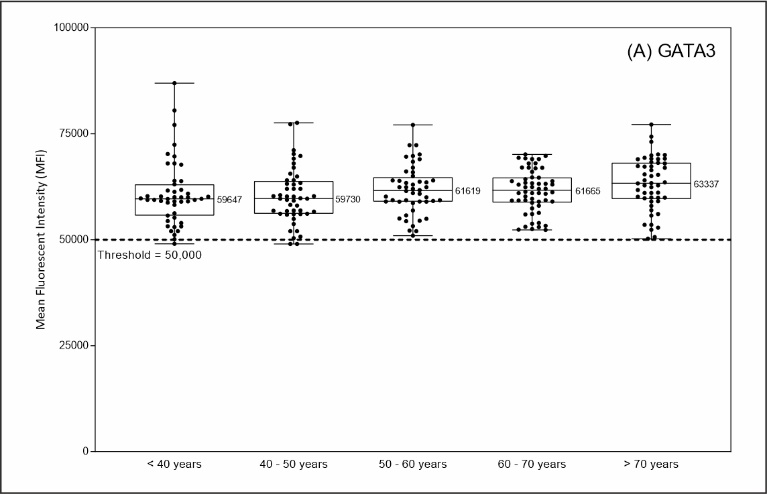

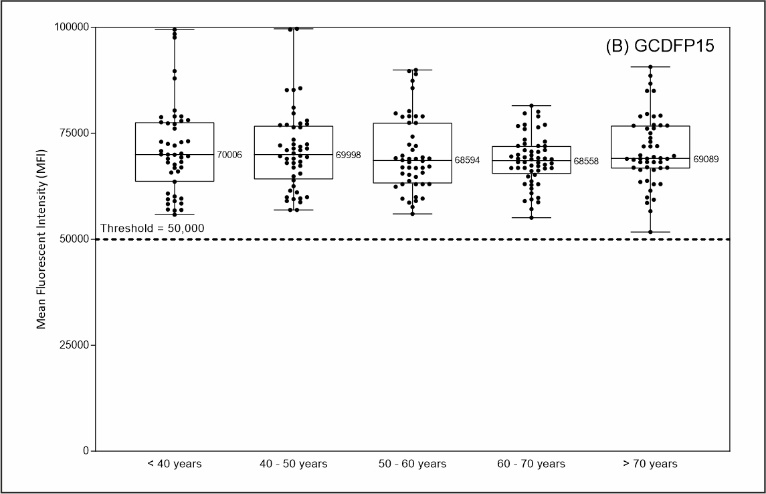

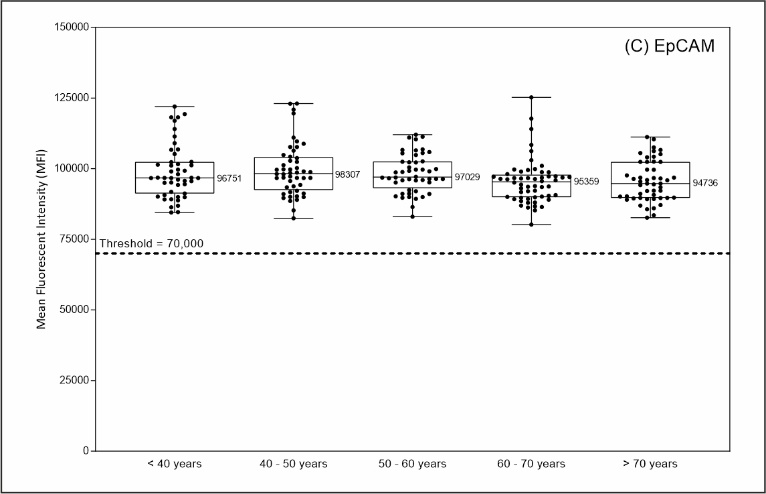

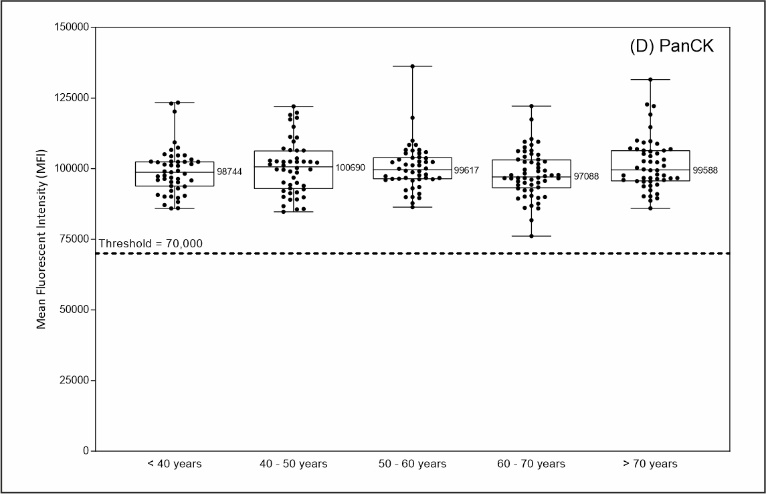

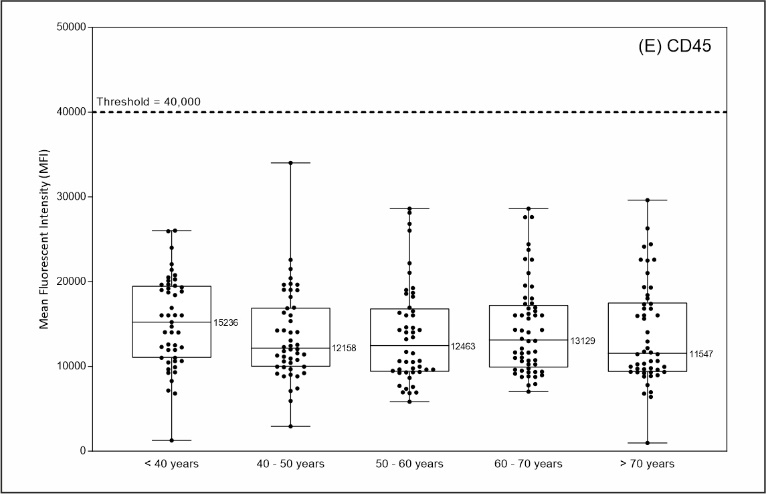

**Supplementary Figure S4. Marker Expression on CTCs from Ductal and Lobular Subtypes.**

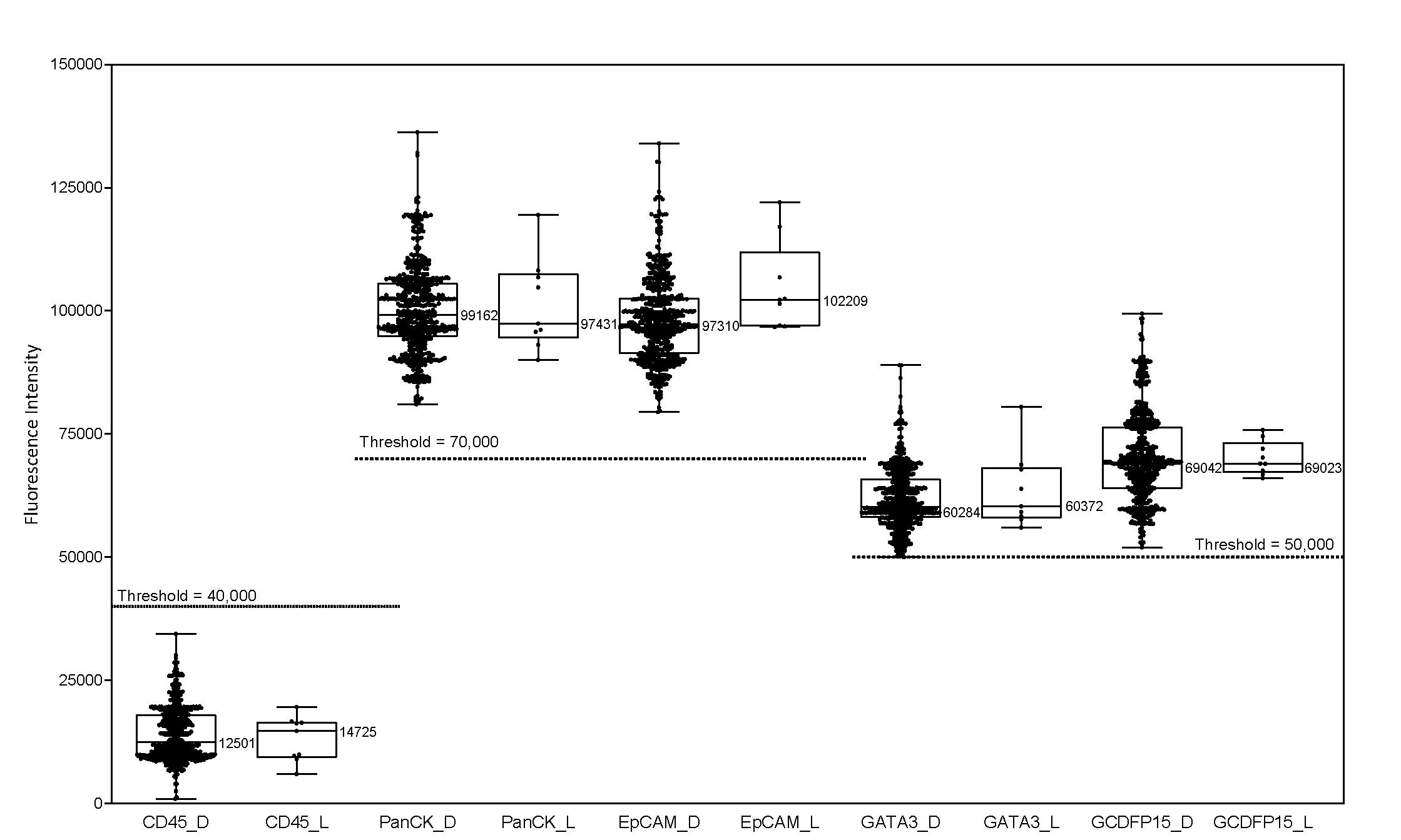

**Supplementary Figure S5. Grade and Marker Expression on CTCs.** (A) GATA3, (B) GCDFP15, (C) EpCAM, (D) PanCK, (E) CD45.

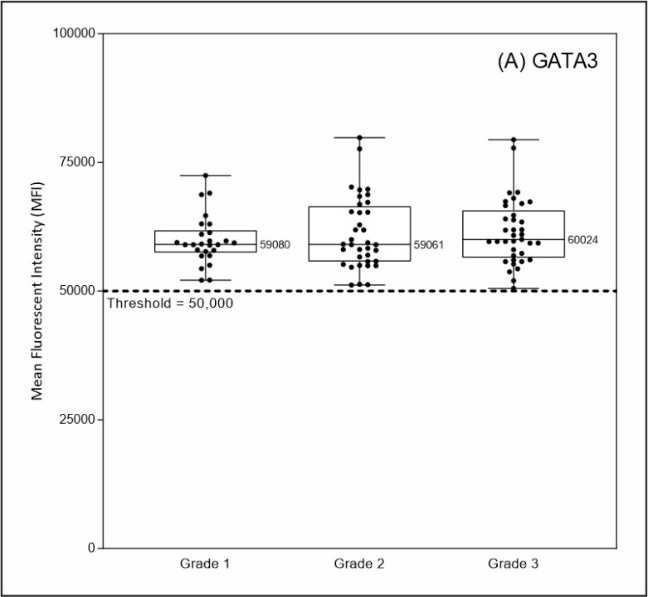

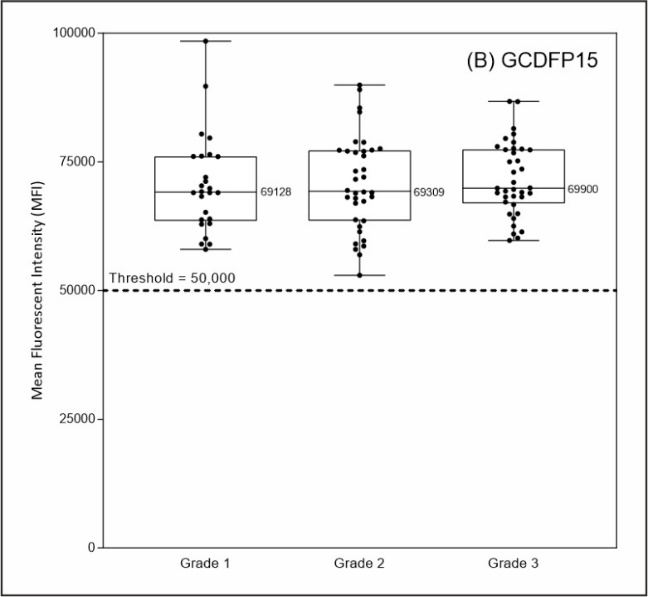

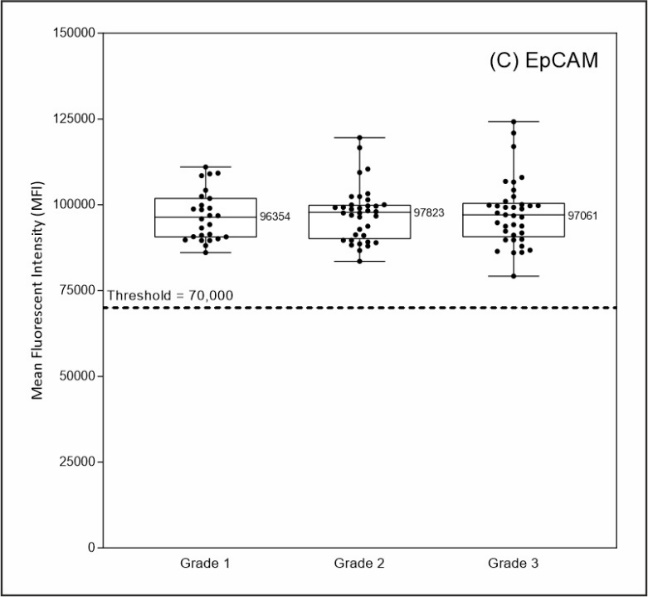

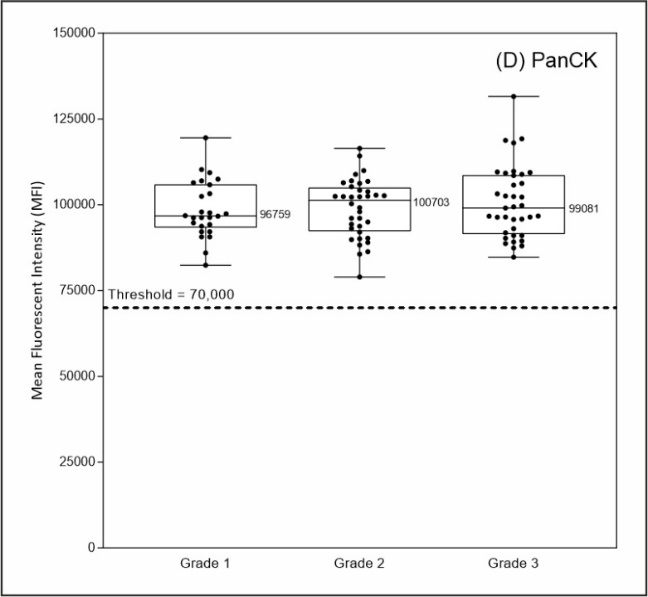

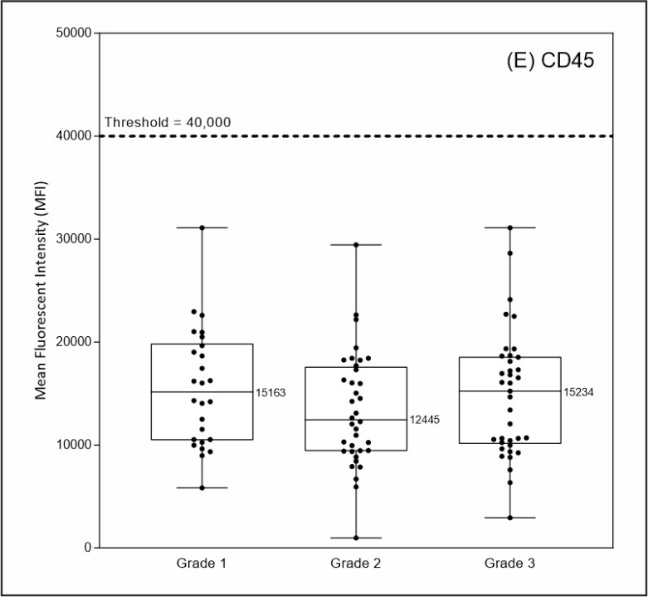

**Supplementary Figure S6. Hormone Receptor Status and Marker Expression on CTCs.** (A) GATA3, (B) GCDFP15, (C) EpCAM, (D) PanCK, (E) CD45.

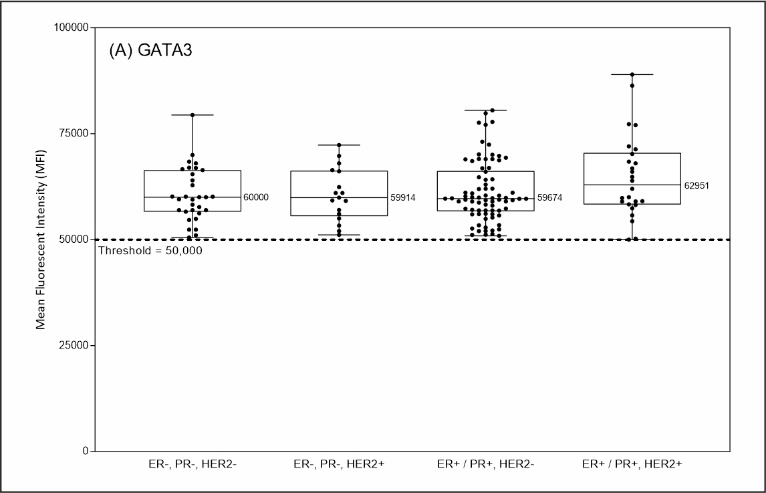

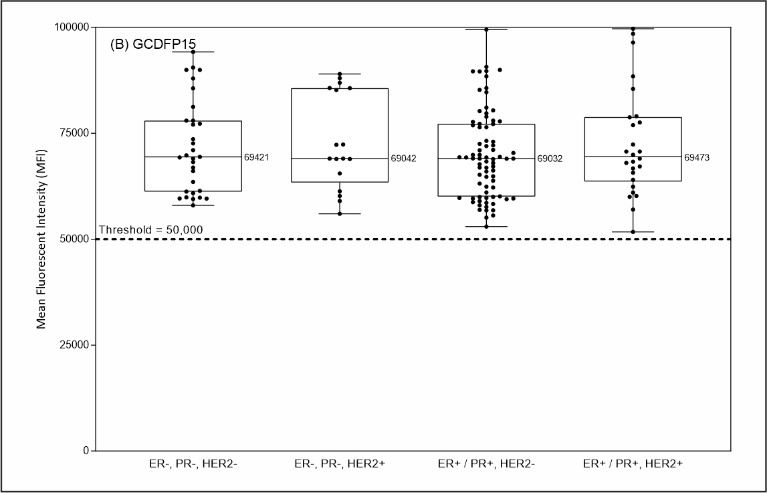

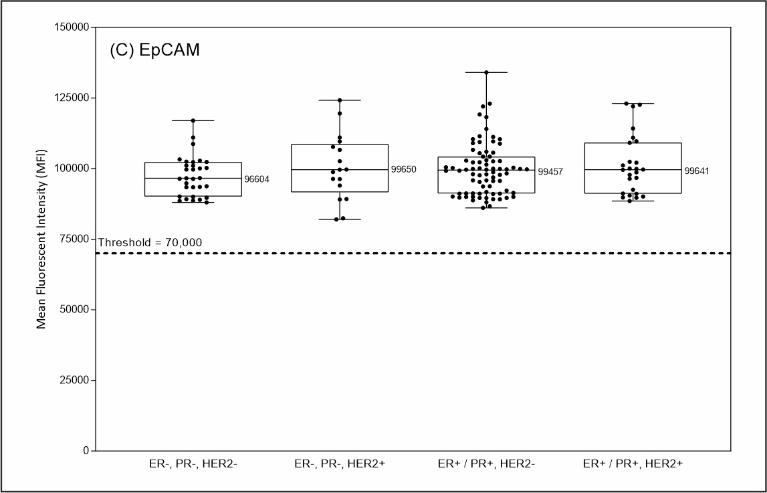

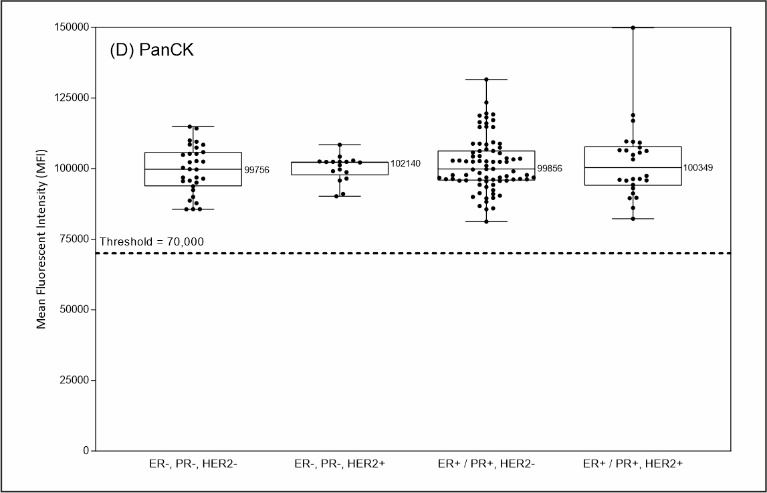

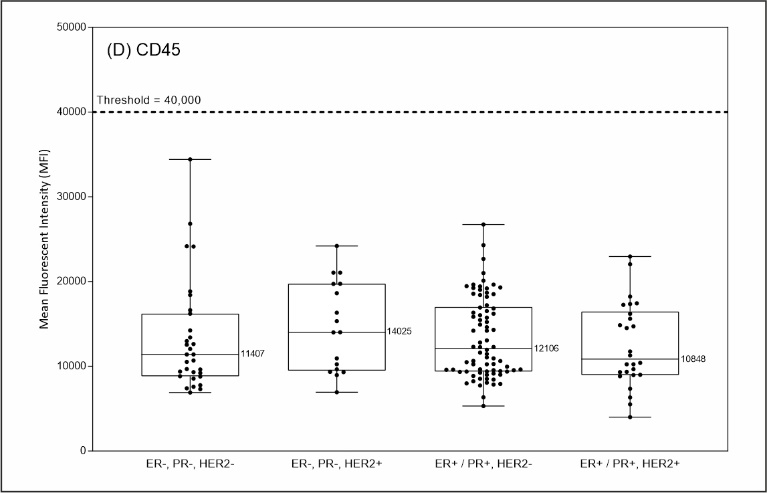

**Supplementary Figure S7. Stage and Marker Expression on CTCs.** (A) GATA3, (B) GCDFP15, (C) EpCAM, (D) PanCK, (E) CD45.

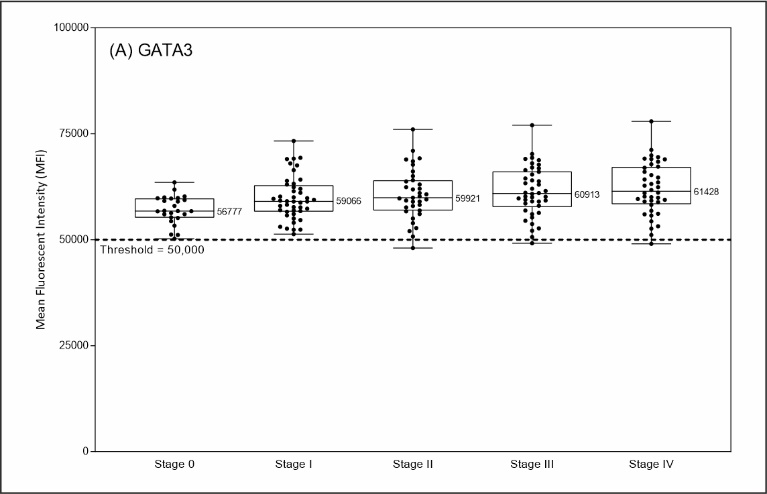

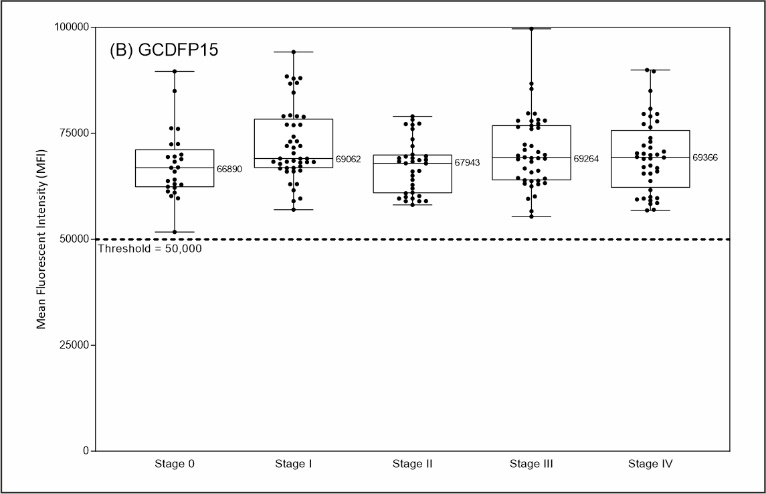

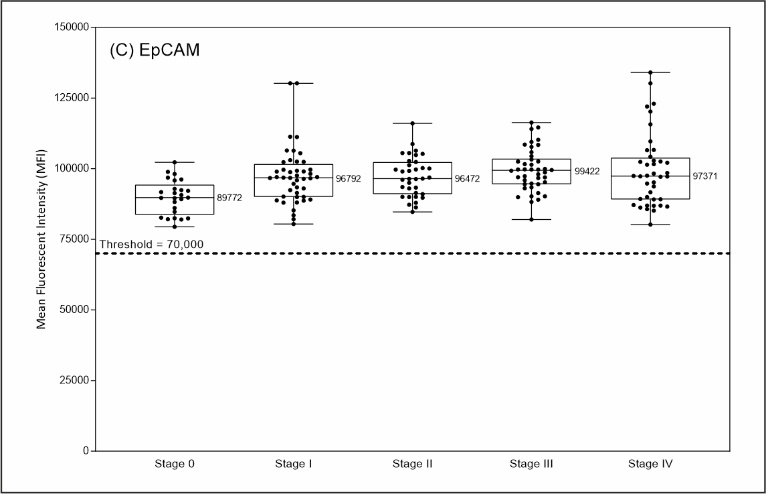

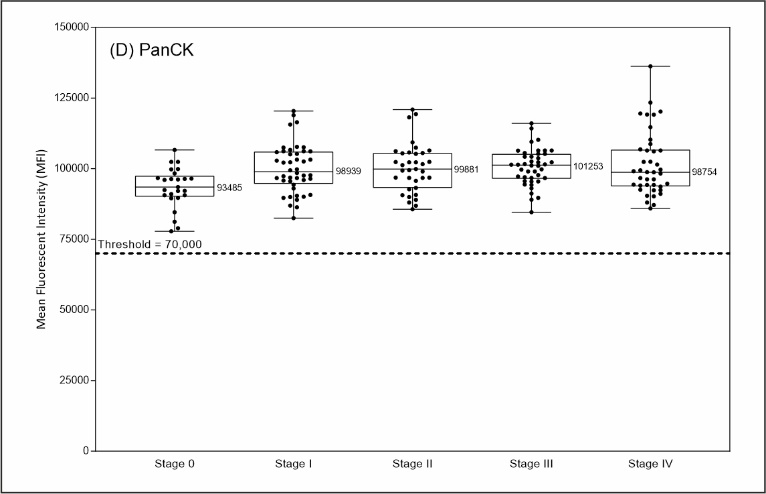

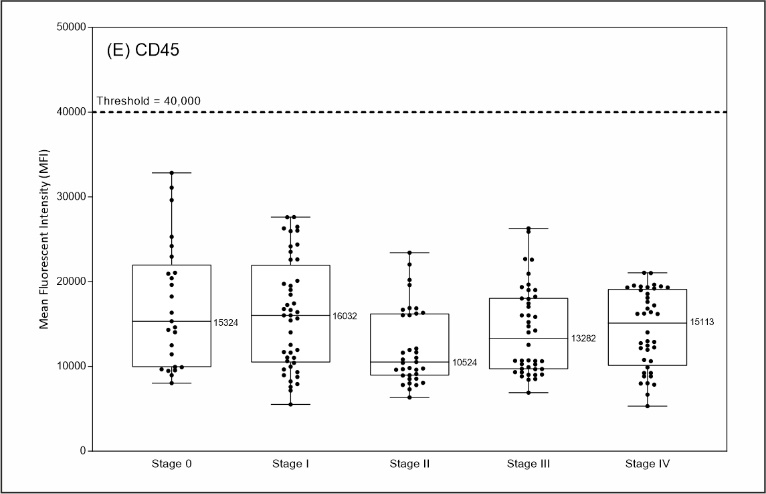

**Supplementary Figure S8. Ethnicity and Marker Expression on CTCs.**

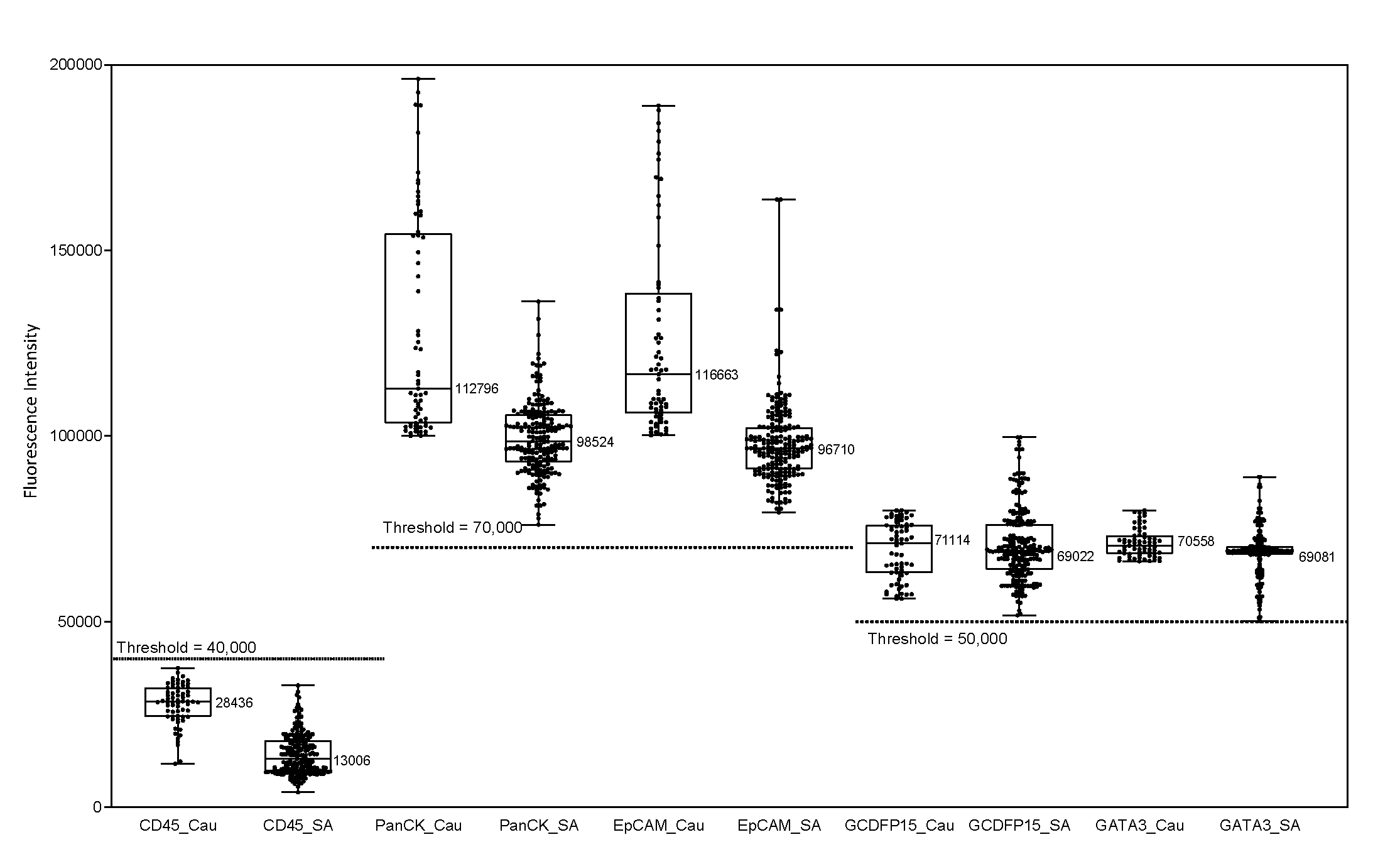

**Supplementary Figure S9: Analytical Validation: Linearity.** The Test exhibited significant linearity with R^2^ ≥0.98. The Tabulated values below the figure show the recovery and range of recovery.

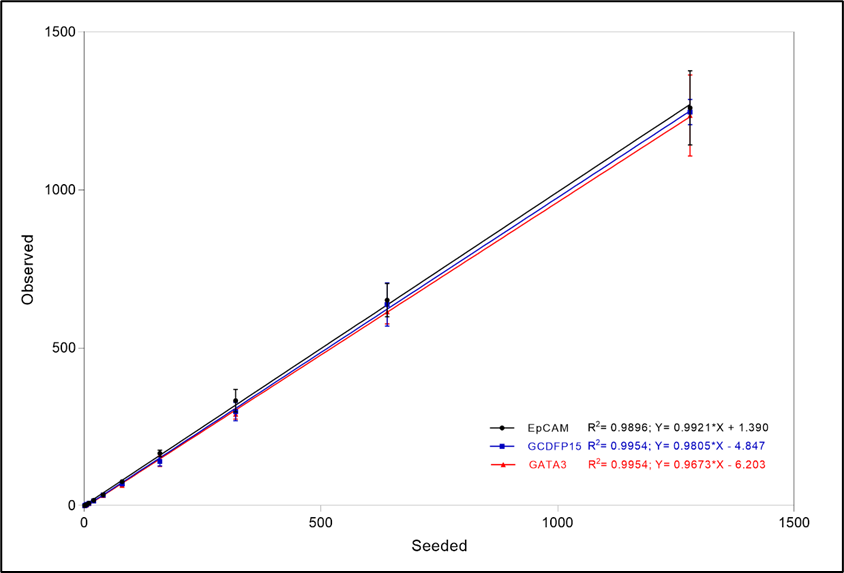

| **Spiked cells** | **Mean and % of Detected Cell Counts (8 Replicates)** | | |
| --- | --- | --- | --- |
|  | **PanCK+,**  **EpCAM+** | **PanCK+,**  **GATA3+** | **PanCK+,**  **GCDFP15+** |
| 1280 | 1260.3 (98.5%)  1125 - 1380 | 1235.8 (96.5%)  1090 – 1473 | 1247.1 (97.4%)  1166 – 1291 |
| 640 | 651.0 (101.7%)  568 – 730 | 614.4 (96.0%)  561 – 689 | 637.3 (99.6%)  531 – 709 |
| 320 | 333.3 (104.1%)  285 – 385 | 290.1 (90.7%)  262 – 310 | 298.8 (93.4%)  250 – 335 |
| 160 | 165.8 (103.6%)  158 – 187 | 141.1 (88.2%)  120 – 168 | 140.6 (87.9%)  128 – 164 |
| 80 | 76.5 (95.6%)  69 – 85 | 64.6 (80.8%)  60 – 70 | 68.5 (85.6%)  62 – 72 |
| 40 | 34.8 (86.9%)  29 – 42 | 32.5 (81.3%)  29 – 34 | 33.6 (84.1%)  30 – 39 |
| 20 | 17.0 (85.0%)  16 – 19 | 16.0 (80.0%)  14 – 17 | 16.0 (80.0%)  14 – 20 |
| 10 | 8.1 (81.3%)  7 – 10 | 8.0 (80.0%)  7 – 9 | 8.3 (82.5%)  7 – 9 |
| 5 | 3.6 (72.5%)  3 – 4 | 3.5 (80.0%)  3 – 4 | 4.3 (85.0%)  3 - 6 |
| 3 | 2.9 (95.8%)  2 – 4 | 1.6 (54.2%)   1 – 2 | 2.0 (66.7%)  1 – 3 |
| 1 | 1.0 (100.0%)  1 - 1 | 0.4 (37.5%)  0 - 1 | 0.3 (25.0%)  0 - 1 |
| 0 | 0 | 0 | 0 |

**SUPPLEMENTARY TABLES**

**Supplementary Table S1. Demographics of Case Control Validation Cohort.**

|  | **Cancer** | **Asymptomatic** |
| --- | --- | --- |
| **N = 10180** | **548** | **9,632** |
| **Age (years)**  Median  Range | 52  21 - 85 | 50  40 - 75 |
| **Cancer Stage**  0  I  II  III  IV | 32  157  158  100  101 | - |

**Supplementary Table S2. Demographics of Prospective Validation Cohort.**

|  | **Cancer** | **Benign** |
| --- | --- | --- |
| **N = 141** | **112** | **29** |
| **Age (years)**  Median  Range | 52  21 - 81 | 39  18 - 69 |
| **Cancer Stage**  0  I  II  III  IV | 24  24  24  20  20 | - |
| **Benign conditions**  Benign fibrocystic disease  Benign parenchyma  Chronic inflammation  Fibroadenoma  Focal duct ectasia  Intraductal papilloma  Reactive lymphoid hyperplasia  Sclerosing adenosis  Focal intraductal hyperplasia | - | 2  8  1  13  1  1  1  1  1 |

**Supplementary Table S3. Validation Set Analysis**

Cancer and asymptomatic samples were randomized into Training and Test Sets in a 70%:30% ratio. Subsequently, all samples were shuffled and random 30% assigned to Test Set 2. The shuffling and 30% randomization was repeated to generate Test Sets 3 - 20. The table reports the findings of the Training and 20 iterations of the Test Sets.

| **Set** | **Sample Type** | **Samples** | **Negative (%)** | **Equivocal (%)** | **Positive (%)** |
| --- | --- | --- | --- | --- | --- |
| **Training** | **Asymptomatic** | **6742** | **6742 (100%)** | **-** | **-** |
|  | **Cancers**  *Stage 0*  *Stage I*  *Stage II*  *Stage III*  *Stage IV* | **384**  *22*  *110*  *111*  *70*  *71* | **30 (7.8%)**  *8 (36.4%)*  *12 (10.9%)*  *9 (8.1%)*  *1 (1.4%)*  *-* | **10 (2.6%)**  *-*  *4 (3.6%)*  *5 (4.5%)*  *1 (1.4%)*  *-* | **344 (89.6%)**  *14 (63.6%)*  *94 (85.5%)*  *97 (87.4%)*  *68 (97.1%)*  *71 (100%)* |
| **Test 1** | **Asymptomatic** | **2890** | **2890 (100%)** | **-** | **-** |
|  | **Cancers**  *Stage 0*  *Stage I*  *Stage II*  *Stage III*  *Stage IV* | **164**  *10*  *47*  *47*  *30*  *30* | **9 (5.5%)**  *2 (20.0%)*  *6 (12.8%)*  *1 (2.1%)*  *-*  *-* | **3 (1.8%)**  *-*  *1 (2.1%)*  *2 (4.3%)*  *-*  *-* | **152 (92.7%)**  *8 (80.0%)*  *40 (85.1%)*  *44 (93.6%)*  *30 (100%)*  *30 (100%)* |
| **Test 2** | **Asymptomatic** | **2890** | **2890 (100%)** | **-** | **-** |
|  | **Cancers**  *Stage 0*  *Stage I*  *Stage II*  *Stage III*  *Stage IV* | **164**  *10*  *47*  *47*  *30*  *30* | **16 (9.8%)**  *3 (30.0%)*  *7 (14.9%)*  *5 (10.6%)*  *1 (3.3%)*  *-* | **5 (3.0%)**  *0 (0.0%)*  *2 (4.3%)*  *3 (6.4%)*  *-*  *-* | **143 (87.2%)**  *7 (70.0%)*  *38 (80.9%)*  *39 (83.0%)*  *29 (96.7%)*  *30 (100%)* |
| **Test 3** | **Asymptomatic** | **2890** | **2890 (100%)** | **-** | **-** |
|  | **Cancers**  *Stage 0*  *Stage I*  *Stage II*  *Stage III*  *Stage IV* | **164**  *10*  *47*  *47*  *30*  *30* | **11 (6.7%)**  *3 (30.0%)*  *5 (10.6%)*  *2 (4.3%)*  *1 (3.3%)*  *-* | **9 (5.5%)**  *-*  *5 (10.6%)*  *4 (8.5%)*  *-*  *-* | **144 (87.8%)**  *7 (70.0%)*  *37 (78.7%)*  *41 (87.2%)*  *29 (96.7%)*  *30 (100%)* |
| **Test 4** | **Asymptomatic** | **2890** | **2890 (100%)** | **-** | **-** |
|  | **Cancers**  *Stage 0*  *Stage I*  *Stage II*  *Stage III*  *Stage IV* | **164**  *10*  *47*  *47*  *30*  *30* | **13 (7.9%)**  *2 (20.0%)*  *9 (19.1%)*  *2 (4.3%)*  *-*  *-* | **3 (1.8%)**  *-*  *2 (4.3%)*  *1 (2.1%)*  *-*  *-* | **148 (90.2%)**  *8 (80.0%)*  *36 (76.6%)*  *44 (93.6%)*  *30 (100%)*  *30 (100%)* |
| **Test 5** | **Asymptomatic** | **2890** | **2890 (100%)** | **-** | **-** |
|  | **Cancers**  *Stage 0*  *Stage I*  *Stage II*  *Stage III*  *Stage IV* | **164**  *10*  *47*  *47*  *30*  *30* | **11 (6.7%)**  *3 (30.0%)*  *6 (12.8%)*  *2 (4.3%)*  *-*  *-* | **3 (1.8%)**  *-*  *1 (2.1%)*  *2 (4.3%)*  *-*  *-* | **150 (91.5%)**  *7 (70.0%)*  *40 (85.1%)*  *43 (91.5%)*  *30 (100%)*  *30 (100%)* |
| **Test 6** | **Asymptomatic** | **2890** | **2890 (100%)** | **-** | **-** |
|  | **Cancers**  *Stage 0*  *Stage I*  *Stage II*  *Stage III*  *Stage IV* | **164**  *10*  *47*  *47*  *30*  *30* | **9 (5.5%)**  *3 (30.0%)*  *4 (8.5%)*  *2 (4.3%)*  *-*  *-* | **5 (3.0%)**  *-*  *4 (8.5%)*  *1 (2.1%)*  *-*  *-* | **150 (91.5%)**  *7 (70.0%)*  *39 (83.0%)*  *44 (93.6%)*  *30 (100%)*  *30 (100%)* |
| **Test 7** | **Asymptomatic** | **2890** | **2890 (100%)** | **-** | **-** |
|  | **Cancers**  *Stage 0*  *Stage I*  *Stage II*  *Stage III*  *Stage IV* | **164**  *10*  *47*  *47*  *30*  *30* | **14 (8.5%)**  *4 (40.0%)*  *6 (12.8%)*  *4 (8.5%)*  *-*  *-* | **4 (2.4%)**  *-*  *1 (2.1%)*  *3 (6.4%)*  *-*  *-* | **146 (89.0%)**  *6 (60.0%)*  *40 (85.1%)*  *40 (85.1%)*  *30 (100%)*  *30 (100%)* |
| **Test 8** | **Asymptomatic** | **2890** | **2890 (100%)** | **-** | **-** |
|  | **Cancers**  *Stage 0*  *Stage I*  *Stage II*  *Stage III*  *Stage IV* | **164**  *10*  *47*  *47*  *30*  *30* | **11 (6.7%)**  *1 (10.0%)*  *5 (10.6%)*  *4 (8.5%)*  *1 (3.3%)*  *-* | **3 (1.8%)**  *-*  *-*  *2 (4.3%)*  *1 (3.3%)*  *-* | **150 (91.5%)**  *9 (90.0%)*  *42 (89.4%)*  *41 (87.2%)*  *28 (93.3%)*  *30 (100%)* |
| **Test 9** | **Asymptomatic** | **2890** | **2890 (100%)** | **-** | **-** |
|  | **Cancers**  *Stage 0*  *Stage I*  *Stage II*  *Stage III*  *Stage IV* | **164**  *10*  *47*  *47*  *30*  *30* | **15 (9.1%)**  *5 (50.0%)*  *7 (14.9%)*  *2 (4.3%)*  *1 (3.3%)*  *-* | **1 (0.6%)**  *-*  *1 (2.1%)*  *-*  *-*  *-* | **148 (90.2%)**  *5 (50.0%)*  *39 (83.0%)*  *45 (95.7%)*  *29 (96.7%)*  *30 (100%)* |
| **Test 10** | **Asymptomatic** | **2890** | **2890 (100%)** | **-** | **-** |
|  | **Cancers**  *Stage 0*  *Stage I*  *Stage II*  *Stage III*  *Stage IV* | **164**  *10*  *47*  *47*  *30*  *30* | **8 (4.9%)**  *2 (20.0%)*  *5 (10.6%)*  *1 (2.1%)*  *-*  *-* | **4 (2.4%)**  *-*  *1 (2.1%)*  *3 (6.4%)*  *-*  *-* | **152 (92.7%)**  *8 (80.0%)*  *41 (87.2%)*  *43 (91.5%)*  *30 (100%)*  *30 (100%)* |
| **Test 11** | **Asymptomatic** | **2890** | **2890 (100%)** | **-** | **-** |
|  | **Cancers**  *Stage 0*  *Stage I*  *Stage II*  *Stage III*  *Stage IV* | **164**  *10*  *47*  *47*  *30*  *30* | **13 (7.9%)**  *2 (20.0%)*  *6 (12.8%)*  *5 (10.6%)*  *-*  *-* | **5 (3.0%)**  *-*  *2 (4.3%)*  *2 (4.3%)*  *1 (3.3%)*  *-* | **146 (89.0%)**  *8 (80.0%)*  *39 (83.0%)*  *40 (85.1%)*  *29 (96.7%)*  *30 (100%)* |
| **Test 12** | **Asymptomatic** | **2890** | **2890 (100%)** | **-** | **-** |
|  | **Cancers**  *Stage 0*  *Stage I*  *Stage II*  *Stage III*  *Stage IV* | **164**  *10*  *47*  *47*  *30*  *30* | **16 (9.8%)**  *4 (40.0%)*  *7 (14.9%)*  *4 (8.5%)*  *1 (3.3%)*  *-* | **3 (1.8%)**  *-*  *1 (2.1%)*  *2 (4.3%)*  *-*  *-* | **145 (88.4%)**  *6 (60.0%)*  *39 (83.0%)*  *41 (87.2%)*  *29 (96.7%)*  *30 (100%)* |
| **Test 13** | **Asymptomatic** | **2890** | **2890 (100%)** | **-** | **-** |
|  | **Cancers**  *Stage 0*  *Stage I*  *Stage II*  *Stage III*  *Stage IV* | **164**  *10*  *47*  *47*  *30*  *30* | **13 (7.9%)**  *2 (20.0%)*  *8 (17.0%)*  *3 (6.4%)*  *-*  *-* | **4 (2.4%)**  *-*  *1 (2.1%)*  *2 (4.3%)*  *1 (3.3%)*  *-* | **147 (89.6%)**  *8 (80.0%)*  *38 (80.9%)*  *42 (89.4%)*  *29 (96.7%)*  *30 (100%)* |
| **Test 14** | **Asymptomatic** | **2890** | **2890 (100%)** | **-** | **-** |
|  | **Cancers**  *Stage 0*  *Stage I*  *Stage II*  *Stage III*  *Stage IV* | **164**  *10*  *47*  *47*  *30*  *30* | **13 (7.9%)**  *3 (30.0%)*  *5 (10.6%)*  *4 (8.5%)*  *1 (3.3%)*  *-* | **2 (1.2%)**  *-*  *-*  *2 (4.3%)*  *-*  *-* | **149 (90.9%)**  *7 (70.0%)*  *42 (89.4%)*  *41 (87.2%)*  *29 (96.7%)*  *30 (100%)* |
| **Test 15** | **Asymptomatic** | **2890** | **2890 (100%)** | **-** | **-** |
|  | **Cancers**  *Stage 0*  *Stage I*  *Stage II*  *Stage III*  *Stage IV* | **164**  *10*  *47*  *47*  *30*  *30* | **15 (9.1%)**  *5 (50.0%)*  *5 (10.6%)*  *5 (10.6%)*  *-*  *-* | **3 (1.8%)**  *-*  *1 (2.1%)*  *1 (2.1%)*  *1 (3.3%)*  *-* | **146 (89.0%)**  *5 (50.0%)*  *41 (87.2%)*  *41 (87.2%)*  *29 (96.7%)*  *30 (100%)* |
| **Test 16** | **Asymptomatic** | **2890** | **2890 (100%)** | **-** | **-** |
|  | **Cancers**  *Stage 0*  *Stage I*  *Stage II*  *Stage III*  *Stage IV* | **164**  *10*  *47*  *47*  *30*  *30* | **11 (6.7%)**  *4 (40.0%)*  *4 (8.5%)*  *2 (4.3%)*  *1 (3.3%)*  *-* | **1 (0.6%)**  *-*  *1 (2.1%)*  *-*  *-*  *-* | **152 (92.7%)**  *6 (60.0%)*  *42 (89.4%)*  *45 (95.7%)*  *29 (96.7%)*  *30 (100%)* |
| **Test 17** | **Asymptomatic** | **2890** | **2890 (100%)** | **-** | **-** |
|  | **Cancers**  *Stage 0*  *Stage I*  *Stage II*  *Stage III*  *Stage IV* | **164**  *10*  *47*  *47*  *30*  *30* | **9 (5.5%)**  *2 (20.0%)*  *4 (8.5%)*  *3 (6.4%)*  *-*  *-* | **4 (2.4%)**  *-*  *1 (2.1%)*  *2 (4.3%)*  *1 (3.3%)*  *-* | **151 (92.1%)**  *8 (80.0%)*  *42 (89.4%)*  *42 (89.4%)*  *29 (96.7%)*  *30 (100%)* |
| **Test 18** | **Asymptomatic** | **2890** | **2890 (100%)** | **-** | **-** |
|  | **Cancers**  *Stage 0*  *Stage I*  *Stage II*  *Stage III*  *Stage IV* | **164**  *10*  *47*  *47*  *30*  *30* | **13 (7.9%)**  *5 (50.0%)*  *7 (14.9%)*  *1 (2.1%)*  *-*  *-* | **2 (1.2%)**  *-*  *1 (2.1%)*  *1 (2.1%)*  *-*  *-* | **149 (90.9%)**  *5 (50.0%)*  *39 (83.0%)*  *45 (95.7%)*  *30 (100%)*  *30 (100%)* |
| **Test 19** | **Asymptomatic** | **2890** | **2890 (100%)** | **-** | **-** |
|  | **Cancers**  *Stage 0*  *Stage I*  *Stage II*  *Stage III*  *Stage IV* | **164**  *10*  *47*  *47*  *30*  *30* | **8 (4.9%)**  *2 (20.0%)*  *5 (10.6%)*  *1 (2.1%)*  *-*  *-* | **4 (2.4%)**  *-*  *-*  *4 (8.5%)*  *-*  *-* | **152 (92.7%)**  *8 (80.0%)*  *42 (89.4%)*  *42 (89.4%)*  *30 (100%)*  *30 (100%)* |
| **Test 20** | **Asymptomatic** | **2890** | **2890 (100%)** | **-** | **-** |
|  | **Cancers**  *Stage 0*  *Stage I*  *Stage II*  *Stage III*  *Stage IV* | **164**  *10*  *47*  *47*  *30*  *30* | **13 (7.9%)**  *4 (40.0%)*  *4 (8.5%)*  *4 (8.5%)*  *1 (3.3%)*  *-* | **4 (2.4%)**  *-*  *2 (4.3%)*  *2 (4.3%)*  *-*  *-* | **147 89.6%)**  *6 (60.0%)*  *41 (87.2%)*  *41 (87.2%)*  *29 (96.7%)*  *30 (100%)* |

**Supplementary Table S4. Expanded findings of the Case Control Clinical Validation Study.** The table below indicates the Median and range of Sensitivities as well as the corresponding Accuracies for cumulative and cancer stage-wise, as observed in the 20-fold cross-validation. 95% confidence interval (CI) are provided for the median values of sensitivity and accuracy. In one analysis for determination of sensitivity and accuracy, samples with equivocal findings were considered as negative and in the other analysis, samples with equivocal findings were considered as positive. Since the test recommends clinical follow-up in individuals with equivocal findings, the final reported values for sensitivity and accuracy (Table 2) are based on samples with equivocal findings being considered as positive. Since none of the samples in the control (cancer-free and asymptomatic) cohort had positive or equivocal findings, overall specificity was 100%.

|  | **Considering Equivocal Findings as Negative** | | **Considering Equivocal Findings as Positive** | |
| --- | --- | --- | --- | --- |
|  | **Sensitivity** | **Accuracy** | **Sensitivity** | **Accuracy** |
| **Cumulative** | Median: 90.55%  Range: 87.20% - 92.68%  95%CI: 89.51% - 91.59% | Median: 99.49%  Range: 99.31% - 99.61%  95%CI: 99.24% - 99.74% | Median: 92.07%  Range: 90.24% - 99.58%  95%CI: 91.12% - 93.03% | Median: 99.57%  Range: 99.48% - 99.74%  95%CI: 99.34% - 99.81% |
| *Stage 0* | Median: 70.00%  Range: 50.00% - 99.98%  95%CI: 34.75% - 93.33% | Median: 99.90%  Range: 99.83% - 99.97%  95%CI: 99.70% - 99.98% | Median: 70.00%  Range: 50.00% - 99.98%  95%CI: 34.75% - 93.33% | Median: 99.90%  Range: 99.83% - 99.97%  95%CI: 99.70% - 99.98% |
| *Stage I* | Median: 85.11%  Range: 76.60% - 89.36%  95%CI: 71.69% - 93.80% | Median: 99.76%  Range: 99.63% - 99.83%  95%CI: 99.51% - 99.90% | Median: 89.36%  Range: 80.85% - 99.82%  95%CI: 76.90% - 96.45% | Median: 99.81%  Range: 99.69% - 99.86%  95%CI: 99.60% - 99.94% |
| *Stage II* | Median: 89.36%  Range: 82.98% - 95.74%  95%CI: 76.90% - 96.45% | Median: 99.83%  Range: 99.73% - 99.93%  95%CI: 99.60% - 99.94% | Median: 95.74%  Range: 89.36% - 99.87%  95%CI: 85.46% - 99.48% | Median: 99.91%  Range: 99.83% - 99.97%  95%CI: 99.75% - 99.99% |
| *Stage III* | Median: 96.67%  Range: 93.33% - 100.0%  95%CI: 82.78% - 99.92% | Median: 99.97%  Range: 99.93% - 100.0%  95%CI: 99.81% - 100.00% | Median: 100.0%  Range: 96.67% - 100.0%  95%CI: 88.43% - 100.00% | Median: 100.0%  Range: 99.97% - 100.0%  95%CI: 99.87% - 100.00% |
| *Stage IV* | Median: 100.0%  Range: 100.0% - 100.0%  95%CI: 88.43% - 100.00% | Median: 100.0%  Range: 100.0% - 100.0%  95%CI: 99.87% - 100.00% | Median: 100.0%  Range: 100.0% - 100.0%  95%CI: 88.43% - 100.00% | Median: 100.0%  Range: 100.0% - 100.0%  95%CI: 99.87% - 100.00% |

**Supplementary Table S5. Prospective Validation Cohort findings**

| **Sample Type** | **Samples** | **Negative (%)** | **Equivocal (%)** | **Positive (%)** |
| --- | --- | --- | --- | --- |
| **Benign** | **29** | **27 (93.1%)** | **2 (6.9%)** | **-** |
| **Cancers**  *Stage 0*  *Stage I*  *Stage II*  *Stage III*  *Stage IV* | **112**  *24*  *24*  *24*  *20*  *20* | **6 (5.4%)**  *3 (2.5%)*  *1 (4.2%)*  *1 (4.2%)*  *1 (5.0%)*  - | -  -  -  -  -  - | **106**  *21 (87.5%)*  *23 (95.8%)*  *23 (95.8%)*  *19 (95.0%)*  *20 (100%)* |

**Supplementary Table S6. Expanded findings of the Prospective Clinical Validation Study.** The table below indicates the stage-wise and cumulative (all stages) sensitivity and accuracy of the test. Among the 141 samples, there were 2 samples with equivocal test findings. Both samples were subsequently diagnosed with benign conditions of the breast. Considering both samples with equivocal findings as negative and as positive, the specificity of the test was 100% and 93.1% respectively (cancer v/s benign). Accuracy was determined based on the differential analysis of samples with equivocal findings.

|  | **Sensitivity** | **Accuracy** | |
| --- | --- | --- | --- |
|  |  | **Equivocal Samples as Negative** | **Equivocal Samples as Positives** |
| **Cumulative** | 94.64%  95% CI: 88.70% - 98.01% | 95.74%  95% CI: 90.97% - 98.42% | 94.33%  95% CI: 89.13% - 97.52% |
| *Stage 0* | 87.50%  95% CI: 67.64% - 97.34% | 94.34%  95% CI: 84.34% - 98.82% | 90.57%  95% CI: 79.34% - 96.87% |
| *Stage I* | 95.83%  95% CI: 78.88% - 99.89% | 98.11%  95% CI: 89.93% - 99.95% | 94.34%  95% CI: 84.34% - 98.82% |
| *Stage II* | 95.83%  95% CI: 78.88% - 99.89% | 98.11%  95% CI: 89.93% - 99.95% | 94.34%  95% CI: 84.34% - 98.82% |
| *Stage III* | 95.00%  95% CI: 75.13% - 99.87% | 97.96%  95% CI: 89.15% - 99.95% | 93.88%  95% CI: 83.13% - 98.72% |
| *Stage IV* | 100.00%  95% CI: 83.16% - 100.00% | 100.00%  95% CI: 92.75% - 100.00% | 95.92%  95% CI: 86.02% - 99.50% |

**Supplementary Table S7. Findings of TaqMan ddPCR Assays.** The table below indicates the various gene variants detected in the tissue samples of 53 breast cancer cases by NGS. ddPCR analysis of these variants in genomic DNA isolated from CTC enriched WBCs, showed an overall 81.1% concordance with findings on tumor tissue.

| **Target Assay** | **Gene ID** | **Control Plasmids** | **Samples** | | |
| --- | --- | --- | --- | --- | --- |
|  |  |  | **Total** | **Positives** | **Negatives** |
| AKT1_E17K | AHWSLXQ | 34R2 | 7 | 5 | 2 |
| EGFR_D855N | AHFBBKR |  | 2 | 2 | 0 |
| ESR1_Y537N | AHCTE56 |  | 3 | 3 | 0 |
| GNAS_R201C | AH6R7EO | 24R3 | 1 | 1 | 0 |
| GNAS_R201H | AH705KW |  | 3 | 2 | 1 |
| KRAS_G12C | AHHS7X9 |  | 1 | 1 | 0 |
| PIK3CA_E545Q | AH21CV0 |  | 1 | 0 | 1 |
| PIK3CA_E542K | AHKA4AQ | 24R5 | 7 | 5 | 2 |
| PIK3CA_E545K | AHLJ2GY |  | 8 | 6 | 2 |
| PIK3CA_H1047L | AHPAWZM | 24R5 | 1 | 1 | 0 |
| PIK3CA_H1047R | AHD2DCF |  | 9 | 8 | 1 |
| PIK3CA_N345K | AHHS7YA |  | 2 | 2 | 0 |
| TP53_R248Q | AHVJNUO | 24R3 | 3 | 3 | 0 |
| TP53_R248W | AHRSTB0 |  | 1 | 0 | 1 |
| TP53_R249S | AHX1J64 |  | 1 | 1 | 0 |
| TP53_R273H | AHUAPOG |  | 2 | 2 | 0 |
| KRAS_G12V | AH0JGKY | Internal | 1 | 1 | 0 |
| **OVERALL** | | | **53** | **43 (81.1%)** | **10** |

**Supplementary Table S8. Demographics of Cohort with Benign or Inflammatory Breast Conditions**

| **Parameter** | **Value** |
| --- | --- |
| **Median Age (Range)** | 34 years (19 – 72 years) |
| **Diagnosis**  Adenosis  Benign Breast Disease  Benign breast parenchyma  Benign Duct Papilloma  Benign Fibrocystic Disease  Benign Fibroepithelial lesion  Benign Phyllodes  Benign Proliferative Disease  Duct Ectasia  Fibroadenoma  Focal intraductal Hyperplasia  Intraductal papilloma  Lactating adenoma  Lipoma  Mastitis  Reactive lymphoid hyperplasia  Squamous metaplasia | 2  13  15  1  2  1  2  2  3  29  3  1  1  1  13  1  1 |

**Supplementary Table S9. Analytical Validation: Stability and Recovery of Spiked Cells.** SKBR3 cells were spiked into healthy donor blood samples and the recovery of spiked cells was evaluated for up to 48 hours.

| **Time (h)** | **Spiked Cells** | **Mean Recovery, % Recovery and Recovery Range (%)** | | |
| --- | --- | --- | --- | --- |
|  |  | **PanCK+,**  **EpCAM+** | **PanCK+,**  **GATA3+** | **PanCK+,**  **GCDFP15+** |
| 0 | 15 | 15.0 (100%)  93.3% - 106.7% | 15.0 (100%)  93.3% - 106.7% | 15 (100%)  93.3% - 106.7% |
| 24 | 15 | 14.7 (97.8%)  93.3% - 100.0% | 14.0 (93.3%)  86.7% - 100.0% | 15 (100%)  93.3% - 106.7% |
| 48 | 15 | 14.7 (97.8%)  93.3% - 100.0% | 13.7 (91.1%)  86.7% - 93.3% | 14.7 (97.8%)  93.3% - 100.0% |

**Supplementary Table S10. Analytical Validation Stability and Recovery of CTCs in Clinical Samples.** Blood samples from known BrAD-CTC positive cases were evaluated for recovery of BrAD CTCs for up to 48 hours.

| **Time (h)** | **Cell Types, Detected Numbers, % Recovery and % Recovery Range** | | | | | | | | |
| --- | --- | --- | --- | --- | --- | --- | --- | --- | --- |
|  | **Patient** | **(a) PanCK+,**  **EpCAM+** | | **(b) PanCK+,**  **GATA3+ cells** | | **(c) PanCK+,**  **GCDFP15+ cells** | | **Total PanCK+ cells (a + b + c)** | |
|  |  | **Cells** | **Recovery** | **Cells** | **Recovery** | **Cells** | **Recovery** | **Cells** | **Recovery** |
| **0 h** | P_1_ | 6 | ***100%*** | 5 | ***100%*** | 6 | ***100%*** | 17 | ***100%*** |
|  | P_2_ | 8 |  | 6 |  | 6 |  | 20 |  |
|  | P_3_ | 6 |  | 6 |  | 7 |  | 19 |  |
|  | P_4_ | 8 |  | 5 |  | 6 |  | 19 |  |
|  | P_5_ | 7 |  | 6 |  | 8 |  | 21 |  |
| **24 h** | P_1_ | 5 | 83.3% | 5 | 100.0% | 5 | 83.3% | 15 | 88.2% |
|  | P_2_ | 7 | 87.5% | 5 | 83.3% | 5 | 83.3% | 17 | 85.0% |
|  | P_3_ | 6 | 100.0% | 5 | 83.3% | 6 | 85.7% | 17 | 89.5% |
|  | P_4_ | 7 | 87.5% | 5 | 100.0% | 6 | 100.0% | 18 | 94.7% |
|  | P_5_ | 7 | 100.0% | 6 | 100.0% | 8 | 100.0% | 21 | 100.0% |
|  | Mean  (Range) | 91.7%  (83.3% - 100.0%) | | 93.3%  (83.3% - 100.0%) | | 90.5%  (83.3% - 100.0%) | | 91.5%  (85.0% - 100.0%) | |
| **48 h** | P_1_ | 5 | 83.3% | 5 | 100.0% | 5 | 83.3% | 15 | 88.2% |
|  | P_2_ | 7 | 87.5% | 5 | 83.3% | 6 | 100.0% | 18 | 90.0% |
|  | P_3_ | 5 | 83.3% | 5 | 83.3% | 6 | 85.7% | 16 | 84.2% |
|  | P_4_ | 7 | 87.5% | 6 | 120.0% | 5 | 83.3% | 18 | 94.7% |
|  | P_5_ | 6 | 85.7% | 5 | 83.3% | 7 | 87.5% | 18 | 85.7% |
|  | Mean  (Range) | 85.5%  (83.3% - 100.0%) | | 94.0%  (83.3% - 120.0%) | | 88.0%  (83.3% - 100.0%) | | 88.6%  (84.2% - 94.7%) | |

**Supplementary Table S11. Analytical Validation: Sensitivity, Specificity, Accuracy.** SKBR3 cells were spiked into healthy donor blood samples at various seed densities and their recoveries evaluated to determine sensitivity. Unspiked healthy donor blood samples were evaluated for false positives to determine specificity. Accuracy was determined from sensitivity and specificity.

| **Spiked** | **Detected Cells: Mean (Range)** | **Negative** | **Positive** |
| --- | --- | --- | --- |
| **PanCK+, EpCAM+, CD45-** | | | |
| **0** | - | 30 | - |
| **5*** | 3.9 (2 – 5) | 2 | 8 |
| **10*** | 9.6 (8 – 10) | - | 10 |
| **20*** | 18.8 (16 – 22) | - | 10 |
| **40*** | 41.4 (34 – 44) | - | 10 |
| **80*** | 76.1 (71 - 80) | - | 10 |
| **PanCK+, GATA3+, CD45-** | | | |
| **0** | - | 30 | - |
| **5*** | 4.4 (3 – 5) | 1 | 9 |
| **10*** | 9.2 (6 – 10) | - | 10 |
| **20*** | 19.7 (16 – 20) | - | 10 |
| **40*** | 40.4 (32 – 40) | - | 10 |
| **80*** | 74.3 (70 - 79) | - | 10 |
| **PanCK+, GCDFP15+, CD45-** | | | |
| **0** | - | 30 | - |
| **5*** | 2.9 (2 – 4) | 3 | 7 |
| **10*** | 6.9 (6 – 7) | - | 10 |
| **20*** | 15.9 (12 – 16) | - | 10 |
| **40*** | 35.7 (30 – 35) | - | 10 |
| **80*** | 65.2 (62 - 71) | - | 10 |
| **Overall PanCK+, CD45-** | | | |
| **0** | - | 30 | - |
| **15** | 11.2 (8 - 13) | 3 | 7 |
| **30** | 25.7 (22 – 26) | - | 10 |
| **60** | 54.4 (46 – 57) | - | 10 |
| **120** | 117.5 (97 – 112) | - | 10 |
| **240** | 215.7 (206 - 228) | - | 10 |
| **represents proportionate number of spiked cells in marker subset analysis* | | | |

**Supplementary Table S12. Analytical Validation: Precision.** Recovery of SKBR3 cells spiked into healthy donor blood samples across multiple replicates by 2 independent operators and over multiple days were used to determine the %CV.

| **A. EpCAM** | **Low Spike (15 cells)** | | | **High Spike (150 cells)** | | | **Overall CV%** |
| --- | --- | --- | --- | --- | --- | --- | --- |
|  | **Mean** | **SD** | **CV%** | **Mean** | **SD** | **CV%** |  |
| **Intra-Run** | | | | | | | |
| User 1 | *15.8* | *1.18* | *7.5%* | *150.4* | *3.18* | *2.1%* | ***4.8%*** |
| User 2 | *15.6* | *1.04* | *6.7%* | *150.6* | *2.83* | *1.9%* | ***4.3%*** |
| Cumulative | *15.7* | *0.96* | *6.1%* | *150.5* | *2.67* | *1.8%* | ***4.0%*** |
| **Inter-Run** | | | | | | | |
| User 1 | *15.8* | *0.58* | *3.7%* | *150.4* | *0.99* | *0.7%* | ***2.2%*** |
| User 2 | *15.6* | *0.53* | *3.4%* | *150.6* | *1.4* | *0.9%* | ***2.2%*** |
| Cumulative | *15.7* | *0.51* | *3.3%* | *150.5* | *0.6* | *0.4%* | ***1.9%*** |
| **Inter-User** | | | | | | | |
| Inter-User | *15.7* | *0.13* | *0.8%* | *150.5* | *0.11* | *0.1%* | ***0.5%*** |
| **OVERALL** | - | - | **7.1%** | - | - | **2.0%** | ***4.6%*** |

| **B. GATA3** | **Low Spike (15 cells)** | | | **High Spike (150 cells)** | | | **Overall CV%** |
| --- | --- | --- | --- | --- | --- | --- | --- |
|  | **Mean** | **SD** | **CV%** | **Mean** | **SD** | **CV%** |  |
| **Intra-Run** | | | | | | | |
| User 1 | 15.6 | 0.95 | ***6.1%*** | 151.6 | 2.45 | ***1.6%*** | ***3.9%*** |
| User 2 | 15.7 | 0.93 | ***5.9%*** | 151.7 | 2.64 | ***1.7%*** | ***3.8%*** |
| Cumulative | 15.6 | 0.85 | ***5.6%*** | 151.6 | 2.34 | ***1.4%*** | ***3.5%*** |
| **Inter-Run** | | | | | | | |
| User 1 | 15.6 | 0.41 | ***2.6%*** | 151.6 | 0.58 | ***0.4%*** | ***1.5%*** |
| User 2 | 15.7 | 0.32 | ***2.0%*** | 151.7 | 0.59 | ***0.4%*** | ***1.2%*** |
| Cumulative | 15.6 | 0.19 | ***1.2%*** | 151.6 | 0.41 | ***0.3%*** | ***0.8%*** |
| **Inter-User** | | | | | | | |
| Inter-User | 15.6 | 0.04 | ***0.2%*** | 151.6 | 0.07 | ***0.0%*** | ***0.1%*** |
| **OVERALL** | **-** | **-** | ***6.0%*** | **-** | **-** | **1.7%** | ***3.9%*** |

| **C. GCDFP15** | **Low Spike (15 cells)** | | | **High Spike (150 cells)** | | | **Overall CV%** |
| --- | --- | --- | --- | --- | --- | --- | --- |
|  | **Mean** | **SD** | **CV%** | **Mean** | **SD** | **CV%** |  |
| **Intra-Run** | | | | | | | |
| User 1 | 15.7 | 0.96 | ***6.1%*** | 150.6 | 2.11 | ***1.4%*** | ***3.8%*** |
| User 2 | 15.7 | 0.96 | ***6.1%*** | 151.3 | 2.25 | ***1.5%*** | ***3.8%*** |
| Cumulative | 15.7 | 0.87 | ***5.5%*** | 150.9 | 2.04 | ***1.5%*** | ***3.5%*** |
| **Inter-Run** | | | | | | | |
| User 1 | 15.7 | 0.43 | ***2.7%*** | 150.6 | 0.59 | ***0.4%*** | ***1.6%*** |
| User 2 | 15.7 | 0.29 | ***1.9%*** | 151.3 | 0.28 | ***0.2%*** | ***1.1%*** |
| Cumulative | 15.7 | 0.31 | ***2.0%*** | 150.9 | 0.38 | ***0.3%*** | ***1.2%*** |
| **Inter-User** | | | | | | | |
| Inter-User | 15.7 | 0.03 | ***0.2%*** | 150.9 | 0.49 | ***0.3%*** | ***0.3%*** |
| **OVERALL** | - | - | ***6.1%*** | - | - | ***1.5%*** | ***3.8%*** |

**Supplementary Table S13. Analytical Validation: Guard Banding Studies for Robustness.** Guard banding studies established the ability of the Test to not be prone to variations from deliberate controlled variations

| **Parameter** | **Range** | | **% Variance** | |
| --- | --- | --- | --- | --- |
|  | **Normal** | **Guard Band**  **(Low, High)** | **Low** | **High** |
| **Blood Collection** | | | | |
| EDTA Vacutainer | Current | New, Near Expiry | 4.9% | 5.1% |
| **RBC Lysis** | | | | |
| Lysis Buffer | 5 vol | 4 vol, 6 vol | 5.9% | 4.9% |
| Incubation Temperature | 37°C | 35°C, 39°C | 2.5% | 3.1% |
| Incubation Time | 20 min | 15 min, 25 min | 2.9% | 2.6% |
| pH | 7.2 | pH 6.7, pH 7.7 | 2.1% | 1.5% |
| Centrifugation Speed | 400 x *g* | 320 x *g,* 480 x *g* | 3.1% | 2.4% |
| Centrifugation Temperature | 4°C | 3.2°C, 4.8°C | 2.5% | 1.5% |
| Centrifugation Time | 5 min | 4 min, 6 min | 4.8% | 4.1% |
| **Immunocytochemistry** | | | | |
| Paraformaldehyde | 4% | 3.5%, 4.5% | 0.9% | 2.8% |
|  | 20 min | 15 min, 25 min | 1.3% | 2.6% |
| Triton X-100 | 0.3% | 0.25%, 0.35% | 2.0% | 2.5% |
|  | 25 min | 20 min, 30 min | 3.5% | 1.9% |
| 1° Ab Dilution: Anti-EpCAM | 1:500 | 1:450, 1:550 | 5.4% | 6.7% |
| 1° Ab Dilution: Anti-PanCK | 1:500 | 1:450, 1:550 | 1.9% | 1.7% |
| 1° Ab Dilution: Anti-CD45 | 1:500 | 1:450, 1:550 | 2.1% | 3.5% |
| 1° Ab Dilution: Anti-GATA3 | 1:4 | 1:3, 1: 5 | 5.9% | 6.6% |
| 1° Ab Dilution: Anti-GCDFP15 | 1:2 | 1:1, 1:3 | 5.3% | 4.4% |
| 1° Ab incubation temperature* | 25°C | 23°C, 27°C | 3.3% | 2.0% |
| 1° Ab Incubation Time* | 60 min | 50 min, 70 min | 2.4% | 3.3% |
| 2° Ab Dilution* | 1: 100 | 1:50, 1: 150 | 2.5% | 2.5% |
| 2° Ab incubation temperature* | 25°C | 23°C, 27°C | 2.6% | 2.0% |
| 2° Ab incubation time* | 60 min | 50 min, 70 min | 1.9% | 3.7% |
| **evaluated for GATA3.* | | | | |

**Supplementary Table S14. Analytical Validation: Impact of Potentially Interfering Substances.** The Test was not prone to interference from endogenous agents (deranged serum parameters) and exogenous agents (common non-anticancer drugs)

| **Agent** | **Concentration Used** | **Detected Cells / mL** | | |
| --- | --- | --- | --- | --- |
|  |  | **PanCK+,**  **EpCAM+** | **PanCK+,**  **GCDFP15+** | **PanCK+,**  **GATA3** |
| Levothyroxine | 140 ng / mL | 9 (90%) | 10 (100%) | 9 (90%) |
| Lisinopril | 58 ng / mL | 9 (90%) | 9 (90%) | 9 (90%) |
| Atorvastatin | 30 ng / mL | 10 (100%) | 9 (90%) | 8 (80%) |
| Metformin | 5 μg / mL | 10 (100%) | 10 (100%) | 10 (100%) |
| Amlodipine | 5 ng / mL | 9 (90%) | 9 (90%) | 9 (90%) |
| Metoprolol | 50 ng / mL | 8 (80%) | 10 (100%) | 10 (100%) |
| Omeprazole | 660 ng / mL | 9 (90%) | 10 (100%) | 10 (100%) |
| Albuterol | 4.2 ng / mL | 9 (90%) | 10 (100%) | 9 (90%) |
| Ranitidine | 450 ng / mL | 9 (90%) | 9 (90%) | 10 (100%) |
| Azithromycin | 500 ng / mL | 9 (90%) | 10 (100%) | 10 (100%) |
| Paracetamol | 9.9 μg / mL | 9 (90%) | 9 (90%) | 9 (90%) |
| Aspirin | 3 μg / mL | 10 (100%) | 10 (100%) | 9 (90%) |
| Loperamide | 3.4 ng / mL | 9 (90%) | 8 (80%) | 10 (100%) |
| Dextromethorphan | 2.9 ng / mL | 10 (100%) | 9 (90%) | 10 (100%) |
| Ulipristal acetate | 170 ng / mL | 9 (90%) | 10 (100%) | 9 (90%) |
| Cholesterol | 3 mg / mL | 9 (90%) | 8 (80%) | 8 (80%) |
| Creatinine | 20 μg / mL | 8 (80%) | 9 (90%) | 8 (80%) |
| Uric Acid | 150 μg / mL | 9 (90%) | 8 (80%) | 8 (80%) |
| Bilirubin | 20 μg / mL | 9 (90%) | 8 (80%) | 8 (80%) |
| Haemoglobin | 200 mg / mL | 10 (100%) | 9 (90%) | 8 (80%) |
| Glucose | 3 mg / mL | 10 (100%) | 9 (90%) | 9 (90%) |
| Control | - | 9 (90%) | 10 (100%) | 10 (100%) |
